# Different responses to neoadjuvant chemotherapy in urothelial carcinoma molecular subtypes

**DOI:** 10.1101/2021.05.11.21255912

**Authors:** Gottfrid Sjödahl, Johan Abrahamsson, Karin Holmsten, Carina Bernardo, Gunilla Chebil, Pontus Eriksson, Iva Johansson, Petter Kollberg, Claes Lindh, Kristina Lövgren, Nour-al-Dain Marzouka, Hans Olsson, Mattias Höglund, Anders Ullén, Fredrik Liedberg

**Affiliations:** Department of Translational Medicine, Lund University, Malmö and Department of Urology Skåne University Hospital, Malmö, Sweden; Department of Oncology-Pathology, Karolinska Institutet and Department of Oncology, Capio S:t Göran Hospital, Stockholm, Sweden; Division of Oncology and Pathology, Department of Clinical Sciences, Lund University, Lund, Sweden; Department of Pathology and Cytology, Karolinska University Hospital, Stockholm, Sweden; Department of Pathology, Linköping University Hospital; Department of Oncology-Pathology, Karolinska Institutet and Department of Pelvic Cancer, Genitourinary Oncology and Urology unit, Karolinska University Hospital, Stockholm, Sweden

## Abstract

For muscle-invasive bladder cancer (MIBC), there are no tissue biomarkers in clinical use that identify patients sensitive or resistant to neoadjuvant chemotherapy. The present study investigates how molecular subtypes impact pathological response and survival in 149 patients receiving preoperative cisplatin-based chemotherapy. Tumor classification was performed by transcriptomic profiling and by a 13-marker immunostaining panel. Furthermore, we explored differential gene expression and chemotherapy response beyond molecular subtypes. Tumors with Genomically Unstable (GU) and Urothelial-like (Uro) subtypes had higher proportions of pathological response and superior survival outcomes as compared to the Basal-Squamous (Ba/Sq) subtype following neoadjuvant chemotherapy and radical cystectomy. Based on our findings, we suggest that urothelial cancer of the luminal-like GU- and Uro-subtypes are more responsive to cisplatin-based chemotherapy. We also found the gene coding for osteopontin (*SPP1*) to display a subtype-dependent effect on chemotherapy response, confirmed at the protein level by immunohistochemistry. Combined analyses of second-generation, subtype-specific biomarkers may be an additional way forward to develop a more precision-based treatment approach for neoadjuvant chemotherapy in MIBC.

## Introduction

Urothelial carcinoma (UC) of the bladder is the fifth most common cancer in Europe and a cause of significant morbidity and mortality.^1^ Despite being one of few treatment recommendations with the highest level of evidence for muscle invasive bladder cancer (MIBC)^2^ there is an underuse of neoadjuvant chemotherapy prior to radical cystectomy (RC).^3^ Reasons for not applying neoadjuvant chemotherapy are treatment-related toxicity and a modest absolute survival benefit, between 5% and 10%.^4^ There are no biomarkers in clinical use that identify patients sensitive or resistant to neoadjuvant chemotherapy to reduce overtreatment of non-responders. This represents an unmet medical need and a major challenge for the research community.^5^

We have previously proposed a molecular taxonomy for bladder cancer,^6-8^ that together with five other RNA-based classification systems provided the basis for a consensus molecular classification of MIBC.^9^ These subtypes represent discrete phenotypic states and are associated with differential enrichment of genomic alterations with potential clinical significance. Varying response to systemic treatments in molecular subtypes has been suggested by previous studies but with disparate results.^9-14^ In addition, genomic alterations in DNA repair genes such as *ERCC2* mutations^15-16^ and *ATM, RB1* or *FANCC* mutations^17^ have been associated with improved response to neoadjuvant chemotherapy.

In this study, we investigated how molecular subtypes impact pathological response and survival in 149 patients receiving preoperative chemotherapy. We classified tumors using pre-defined Lund taxonomy (LundTax) classification algorithms^18^ by transcriptomic profiling and by applying a 13-marker immunostaining panel.^7^ Furthermore, in exploratory differential gene expression analyses, gene signatures predicting chemotherapy response beyond molecular subtypes were investigated. We identified osteopontin (*SPP1*) expression and the status of the tumor suppressors p16 (*CDKN2A*), Rb (*RB1*), and p53 (*TP53*) as subtype-dependent biomarkers for preoperative chemotherapy response.

## Materials and methods

### Patient cohort, outcome measures and follow-up

All patients receiving preoperative cisplatin-based chemotherapy undergoing RC for MIBC between 2004 and 2015 were retrospectively identified in two population-based areas in Sweden (Stockholm and Southern Health Care Region). Out of 172 patients, six who only received one course of preoperative chemotherapy, one without MIBC, and 16 for whom we could not obtain gene expression data were excluded (Supplementary Figure 1). Thus, the final study cohort consisted of 149 patients (referred to as the Chemo-cohort), of which 125 received neoadjuvant (referred to as Neo-cohort), and 24 induction chemotherapy (referred to as Ind-cohort), i.e. preoperative chemotherapy for node positive disease (n=20) or cT4b tumor (n=4)). The median age of the 116 males and 33 females included in the study was 67 (Inter Quartile Range (IQR) 61-70) years. The clinical stage distribution and node status are given in Table 1. In conjunction with RC, a lymphadenectomy was always performed. In the 24 patients receiving induction chemotherapy, the lymphadenectomy was performed up to the aortic bifurcation, whereas in the 125 clinically node-negative patients receiving neoadjuvant chemotherapy, the extent of the lymphadenectomy was at the discretion of the operating surgeon in the five hospitals where the surgery was performed. The preoperative chemotherapy combinations and number of cycles received are reported in Table 1. Ninety-eight percent of patients were treated either with three courses of a four-week gemcitabine/cisplatin regimen (82/149 patients, 55%) or three courses of dose-dense MVAC (methotrexate/vinblastine/doxorubicin/cisplatin) (64/149, 43%). Among the 24 patients treated in an induction setting, a maximum of six courses were administered with a median of four courses. In the Neo-cohort, 46/125 (37%) died from bladder cancer and the corresponding proportion for the Ind-cohort was 16/24 (67%) (Table 1).

**Table 1.**
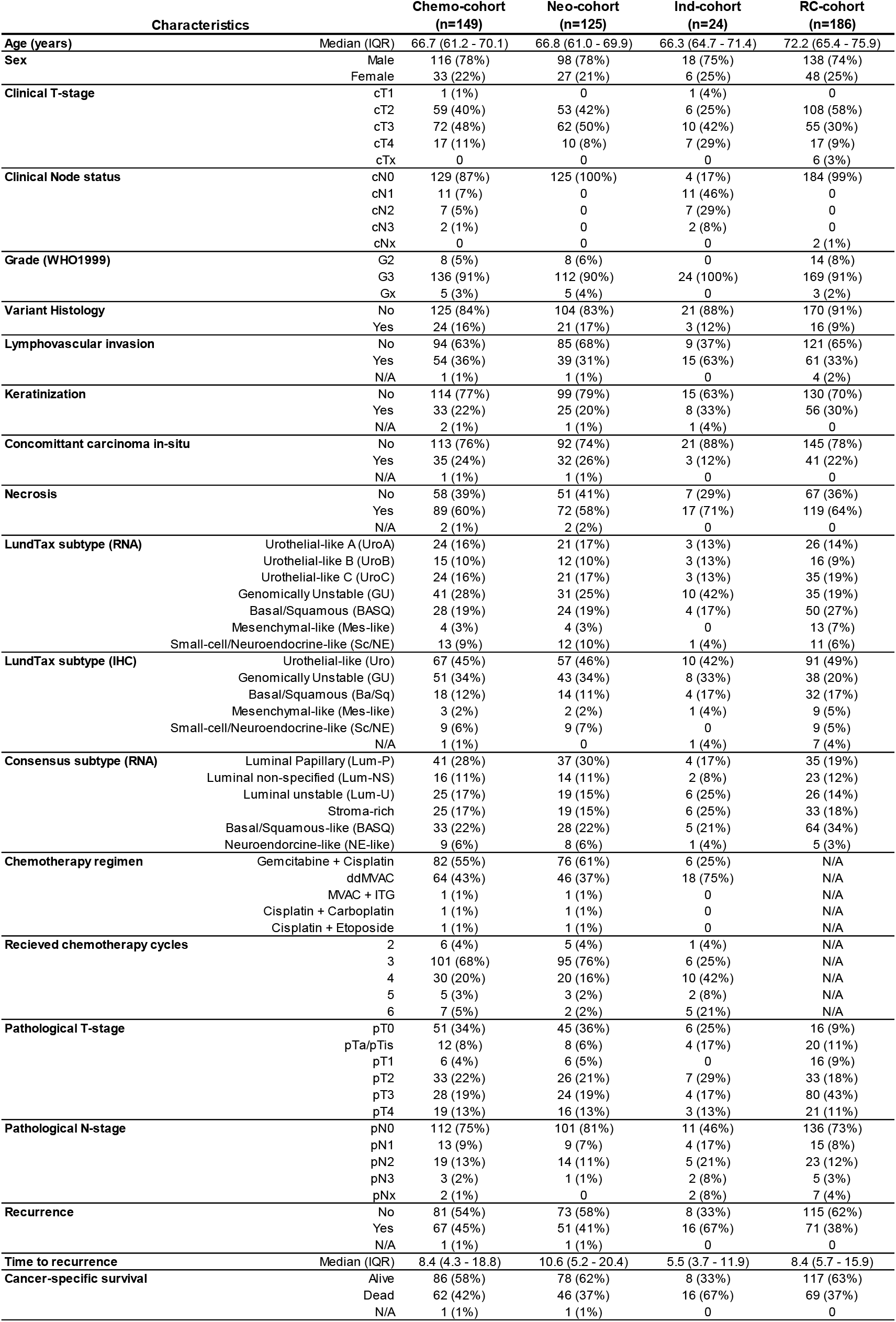
Clinical and pathological characteristics of the study cohorts. IQR, interquartile range; ITG, ifosfamide-paclitaxel-gemcitabine; ddMVAC,dose-dense methotrexate-vinblastine-doxorubicin-cisplatin.

We also established a reference cohort, referred to as the RC-cohort, treated with RC without perioperative chemotherapy in the same hospitals in the Southern Health Care Region between 2006 and 2011. The RC-cohort is based on a published consecutive study^19^ and after exclusions due to RC for non muscle-invasive disease, non-curative RC, clinically node positive disease, and non-urothelial histology, 186 patients remained (Supplementary Figure 1).

The primary outcome measure was pathologic response in the cystectomy specimen, stratified as complete response (pCR, ypT0N0), partial response (pPR, ypT1N0, ypTaN0, or ypTisN0), and no response (yp≥ T2 or ypN+). Pathologic downstaging (pDS) was also stratified as the difference between clinical and pathological stage, and a three-step difference or more was considered a major response.^20^ For the Ind-cohort, pN0 was considered one pDS step. Secondary outcome measures were cancer-specific survival (CSS) and overall survival (OS). CSS and OS were defined as time from RC to death from bladder cancer and death from all causes, respectively. Median follow-up time was 73 (IQR 61-91) months for the Chemo-cohort and 72 (IQR 49-92) months for the RC-cohort as determined by the reverse Kaplan-Meier method. Clinical and pathological data are available in the Supplementary data file.

### Pathological evaluation and immunostaining

Sections from the transurethral resection of the bladder (TUR-B) specimens were re-reviewed by four experienced uropathologists (IJ, CL, HO, GC) to determine T-stage, tumor grade (WHO1999), presence of lymphovascular invasion, keratinization, carcinoma in situ, necrosis, and/or presence of variant histology.^21^ Among variant histologies, there were 16 UCs with divergent differentiation (nine squamous, three glandular, five small-cell or neuroendocrine) and three sarcomatoid, one nested, and one clear-cell variant. Two 1.0 mm cores from representative areas of each TUR-B specimen were assembled into tissue microarrays (TMA), and TMA-sections were stained with 13 antibodies constituting the Lund taxonomy (LundTax) subtyping immunohistochemistry (IHC) marker panel. Antibodies, staining conditions, blinded evaluation, cut-off values and calculation of subtype scores were pre-defined and performed as previously described^7^ (Supplementary Table 1). Anti-osteopontin (*SPP1*) was investigated as a potential chemotherapy response marker and positivity was evaluated semi-quantitatively in cancer cells and in stromal cells separately. Tumor suppressor status was inferred from IHC-data as previously described.^22^ Briefly, the IHC-evaluation for Rb1 was evaluated as percentage of cancer cells with nuclear positivity scored in 10% bins (0-9), where a score ≤ 4 was considered altered. The score for p16 (CDKN2A) was evaluated as staining intensity of cancer cells (0-3), where a score ≤ 0.5 was considered altered. The p53 staining pattern corresponding to genomic alterations (overexpression, complete absence, cytoplasmic) were recorded as altered, whereas wild-type pattern was recorded as non-altered.^23^ Tumors with discordant p53 evaluation were excluded from the analyses. All IHC evaluations were performed blinded to the RNA-classification and study endpoints.

### Transcriptome analysis

The same TUR-B block used for TMA construction was sectioned and stained with hematoxylin and eosin, and tumor rich areas located as close as possible to the positioning of the TMA cores were macrodissected. Tissue from 4-10 10 µm sections of the macrodissected area was used to extract total RNA (HighPure FFPE kit, Roche) as previously described.^7^ Total RNA was amplified, labeled (GeneChip WT Pico Kit, Applied Biosystems) and hybridized to microarrays (Human Gene 1.0 ST array, Affymetrix). Raw data was RNA normalized, adjusted for labeling batch using ComBat^24^ and merged by median probe set values per gene symbol. Classification into the Lund Taxonomy (LundTax) molecular subtypes was performed according to the classification scheme described in Marzouka et al.^8^ Briefly, a k-Top scoring Pairs single-sample molecular subtype predictor was generated using the Bioconductor R package ‘switchBox’ to first predict the 5 main subtypes (Urothelial-like, Uro; Genomically Unstable, GU; Basal-Squamous, Ba/Sq; Mesenchymal-like, Mes-like; Small Cell/Neuroendocrine-like, Sc/NE).^18^ Tumors classified as Uro were then further sub-classified with a second similar classifier into UroA, UroB, and UroC subsets. The LundTax subtype classifier was applied to RMA-normalized, batch-adjusted, quantile normalized, uncentered expression data for each included tumor sample. Raw and normalized data are deposited in the Gene Expression Omnibus under accession number GSE169455.

### Statistical analyses

Two sided χ2-test without continuity correction was used to test associations between categorical variables. If the expected frequency was below 5 for any cell, p-values were calculated based on simulation. Ordinal categorical data were tested by ordered χ2-test (asymptotic linear-by-linear test) and associations with continuous data were tested by Mann-Whitney U test. Multivariable logistic regression models using pCR as dependent variable were constructed for each cohort separately. Kaplan-Meier curves and Cox-proportional hazards (CoxPH) models were used to investigate the association between molecular subtype and survival endpoints.

Genes associated with pCR were identified using moderated t test (‘limma’ R package) and significance analysis of microarrays (SAM). Adjusted p-(‘limma’) or q-values (SAM) below 0.05 were considered significant. Gene set enrichment analysis^25^ used the GSEA software v4.0.3 and a pre-ranked (association to pCR by the ‘limma’ moderated t statistic) gene list with FDR-adjustment for multiple testing. Unsupervised gene sets scanned included MsigDB v7.1 (H. Hallmark, C2 Curated, and C6 Oncogenic). Supervised signatures (n=22) added include the Lund-set of coherent biology signatures in bladder cancer, one-versus rest signatures for LundTax subtypes, and proposed external chemotherapy response signatures split into up- and down-regulated components (n=5) (Supplementary Table 2). Mean values of signatures were used to plot ROC curves and calculate area under the curve using the ‘ROCR’ R package.

### External gene signatures and data sets

Three published gene expression signatures predicting chemotherapy response were identified in the literature, one consisting of genes downregulated in responders,^26^ and two that comprised of both up- and down-regulated genes.^27-28^ We matched as many of the gene symbols as possible to our data set (Supplementary Table 2). For the bidirectional signatures we derived a ratio of mean values of up- and down-signatures. Gene expression data generated by Seiler and colleagues^12^ was obtained via Gene Expression Omnibus accession number GSE87304. Patients were selected by the same inclusion criteria as the Neo-cohort, thereby excluding non-cisplatin containing regimens, patients who received only one chemotherapy cycle, patients with incomplete stage information, and clinically stage T4b/N+ patients. Raw CEL-files were downloaded and processed as described above for the current cohort. The normalized gene expression matrix and clinical response data for the study by Taber and colleagues was retrieved from the supplementary data.^10^ Of the 96 patients used, 21 were treated with neoadjuvant chemotherapy and 75 were treated with first line cisplatin-based chemotherapy in the metastatic setting. Single-sample LundTax classification of tumor samples from external cohorts (Seiler, n=190 and Taber, n=96) was performed on normalized but uncentered expression data.

The study was approved by the Research Ethics Board of Lund University (2012/22; 2013/264; 2017/37)

## Results

### Molecular classification in patients treated with preoperative chemotherapy

We applied molecular subtype classification to tumors from 149 patients treated with preoperative chemotherapy. All patients treated with preoperative chemotherapy are referred to as the Chemo-cohort (n=149). Among these patients, a subset received neoadjuvant treatment (Neo-cohort, n=125) and a subset with nodal spread or clinical stage T4b received induction treatment (Ind-cohort, n=24). We also established a reference cohort not receiving chemotherapy (RC-cohort, n=186). The LundTax molecular subtype proportions arrived at by RNA- and IHC-based classification are illustrated in Figure 1A. The five-group concordance between RNA- and IHC-subtypes in the Chemo-cohort was 0.66 and the overlap between the LundTax and the Consensus-classification systems was as expected (Supplementary Figure 2). The GU subtype was overrepresented among patients treated in an induction setting (Supplementary Figure 3) but there was no significant difference in subtype distribution between the Neo- and RC-cohorts (5×2 χ^2^-test, p=0.06). Hematoxylin and eosin staining, and subtype-defining immunostainings are shown in Figure 1B for histologically representative specimens of the five main LundTax subtypes; Urothelial-like (Uro, further subdivided into UroA, UroB, and UroC), Genomically Unstable (GU), Basal-Squamous (Ba/Sq), Mesenchymal-like (Mes-like) and Small Cell/Neuroendocrine-like (Sc/NE).

**Figure 1.**
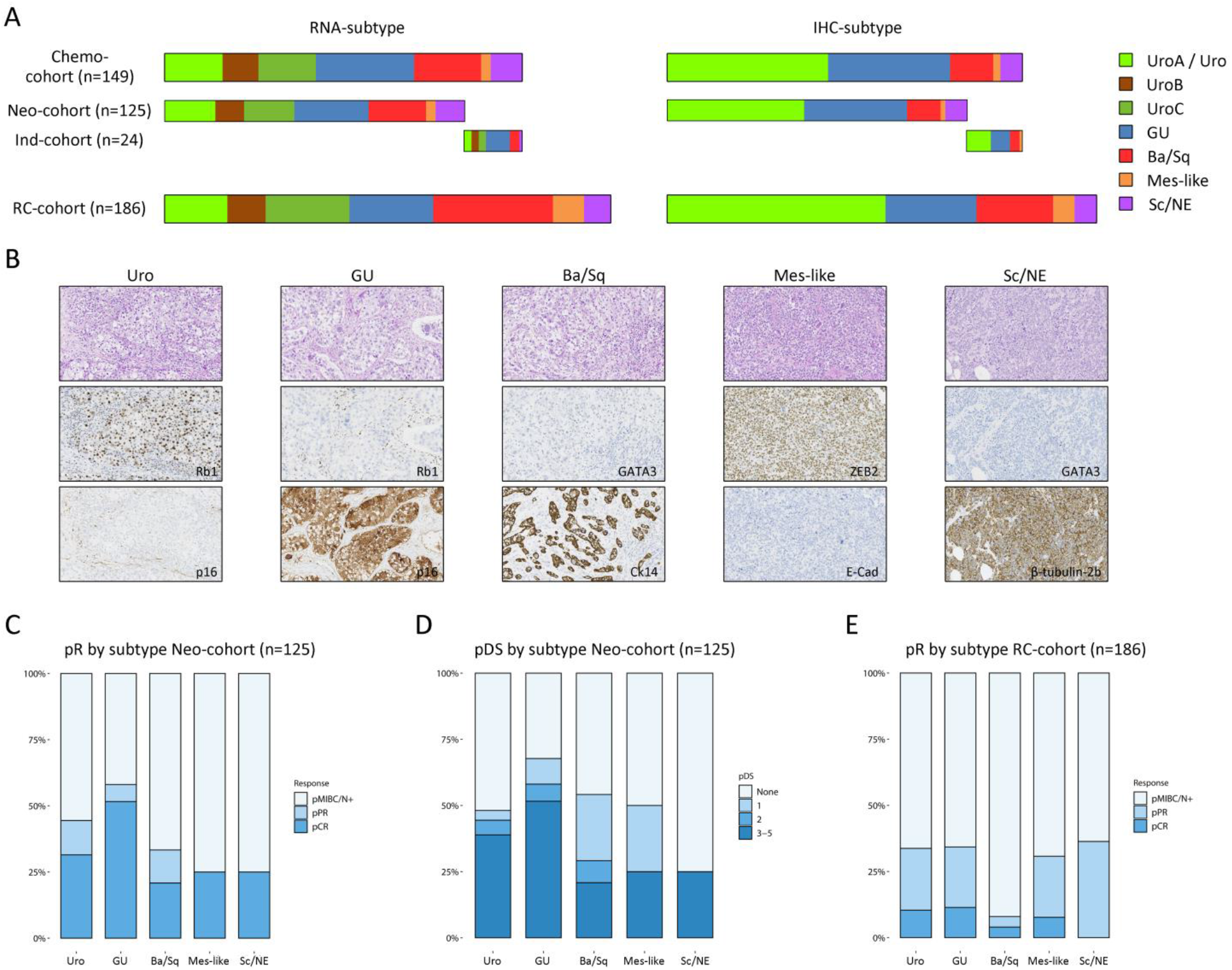
RNA-based molecular subtypes are associated with pathologic response to neoadjuvant chemotherapy. Bar lengths in A) are proportional to the sample size in the Chemo-cohort, the subsets treated with neoadjuvant (Neo-cohort) and induction chemotherapy (Ind-cohort), and the reference cohort treated only with radical cystectomy (RC-cohort). In the left panel, the bars are color coded by RNA-based, and in the right panel by IHC-based LundTax subtypes. The Urothelial-like (Uro) subtype can be further sub-divided into UroA, UroB, and UroC only by RNA-subtyping. Images in B) show Hematoxylin and Eosin-(top), and two subtype defining immunostainings (middle and bottom) for one representative tumor (0.6 x 0.35 mm) from each of the five major molecular subtypes. Stacked barplots in C) display the proportion achieving pathologic complete response (pCR), pathologic partial response (pPR), or having residual muscle-invasive or node positive disease (pMIBC/N+) in the Neo-cohort stratified by RNA-based classification into the 5 major molecular subtypes. Stacked barplots in D) show the number of pathologic downstaging steps achieved by the 5 major molecular subtypes in the Neo-cohort, and in E) the corresponding pathologic response proportions in the RC-cohort by RNA-based molecular subtypes.

### Molecular subtypes are associated with pathologic response in the neoadjuvant setting

The proportion with pathologic complete response (pCR (ypT0N0)) in the Chemo-cohort was 32% and an additional 9% had a pathologic partial response (pPR). Stratification into LundTax-RNA subtypes revealed highest proportion pCR in the GU (41%) and Uro (32%) subtypes and lowest in the Ba/Sq subtype (21%) (p=0.08) (Supplementary Figure 4). The rare Mes-like and Sc/NE subtypes responded similar to Ba/Sq tumors, but the limited number of patients makes interpretation for these subtypes uncertain. Since a complete response is more difficult to obtain for patients with cT4b and/or nodal metastases (Ind-Cohort) and as these patients had different subtype proportions (Supplementary Figure 3), we henceforth analyzed chemotherapy response in the Neo-cohort. Among these patients we observed significantly different proportions of pCR between GU (52%) and Ba/Sq (21%) subtypes (p=0.02) (Figure 1C). This difference was validated/confirmed when analyzing stepwise pathologic downstaging (pDS) and stratifying patients by major response defined as three or more downstaging steps^20^ (52% vs 21%, p=0.02) (Figure 1D). Following the GU subtype, the Uro subtype had the second highest proportion pCR (31%). When further sub-classifying Uro tumors, the proportion with pCR was high in UroA (43%) but lower in UroB (25%), and UroC (24%) tumors suggesting a possible differential response also within the Uro subtype. In a multivariable logistic regression including molecular subtypes and clinical stage (Supplementary Table 3), the odds ratio (OR) for pCR was 0.19 (95% CI: 0.08 - 0.44) for clinical stage group cT3-T4a compared to cT2 and 3.46 (95% CI: 0.95 - 12.59) for GU tumors with Ba/Sq tumors as reference.

The proportion of patients achieving pCR in the RC-cohort was 8% (15/186); 11% (4/35) for GU, 10% (8/77) for Uro, and 4% (2/50) for Ba/Sq tumors (Figure 1E). Among GU and Uro tumors, 34% of cases (12/35 for GU and 26/77 for Uro) displayed a pCR or pPR compared to 8% (4/50) of the Ba/Sq tumors (p=0.002).

### Molecular subtypes are associated with survival independently of tumor stage in the neoadjuvant setting

In the Neo-cohort, patients with Ba/Sq and Sc/NE tumor subtypes had worse cancer-specific survival (CSS) compared to patients with GU and Uro subtypes (Figure 2A), and CSS was also inferior for patients with higher clinical tumor stage (Figure 2B). In a univariable Cox proportional-hazards (CoxPH) regression model, CSS was associated with GU subtype with a hazard ratio (HR) of 0.29 (95% CI: 0.11 - 0.76) with Ba/Sq as reference. Clinical stage group cT3-T4a obtained a HR of 4.91 (95% CI: 2.29 - 10.53) with cT2 as reference (Supplementary Table 4). The 5-year cumulative incidence of recurrence was 27% for the GU subtype in the Neo-cohort compared to 42% in the RC-cohort, while for Ba/Sq tumors the corresponding rates were 52% and 50%, respectively (Supplementary Figure 5). In a multivariable CoxPH regression model adjusting for clinical stage, GU was associated with CSS (HR 0.29, 95% CI: 0.11 - 0.79, p=0.015). Furthermore, the UroC subtype (HR 0.37, 95% CI: 0.14 - 0.94, p=0.036) and clinical stage cT3-T4a were also associated with CSS (HR 5.76, 95% CI: 2.63 - 12.59, p=1.2E-05) (Table 2). The same multivariable CoxPH model for overall survival (OS) resulted in HRs 0.41 (95% CI: 0.17 - 0.96, p=0.04) for the GU subtype, 0.44 (95% CI: 0.19 - 1.04, p=0.06) for the UroC subtype, and 5.39 (95% CI: 2.66 - 10.9, p=2.8E-06) for clinical stage group cT3-T4a (Supplementary Table 5). A similar analysis performed in the RC-cohort showed no associations between CSS and molecular subtype or clinical stage (Supplementary Figure 6 and Supplementary Table 6).

**Figure 2.**
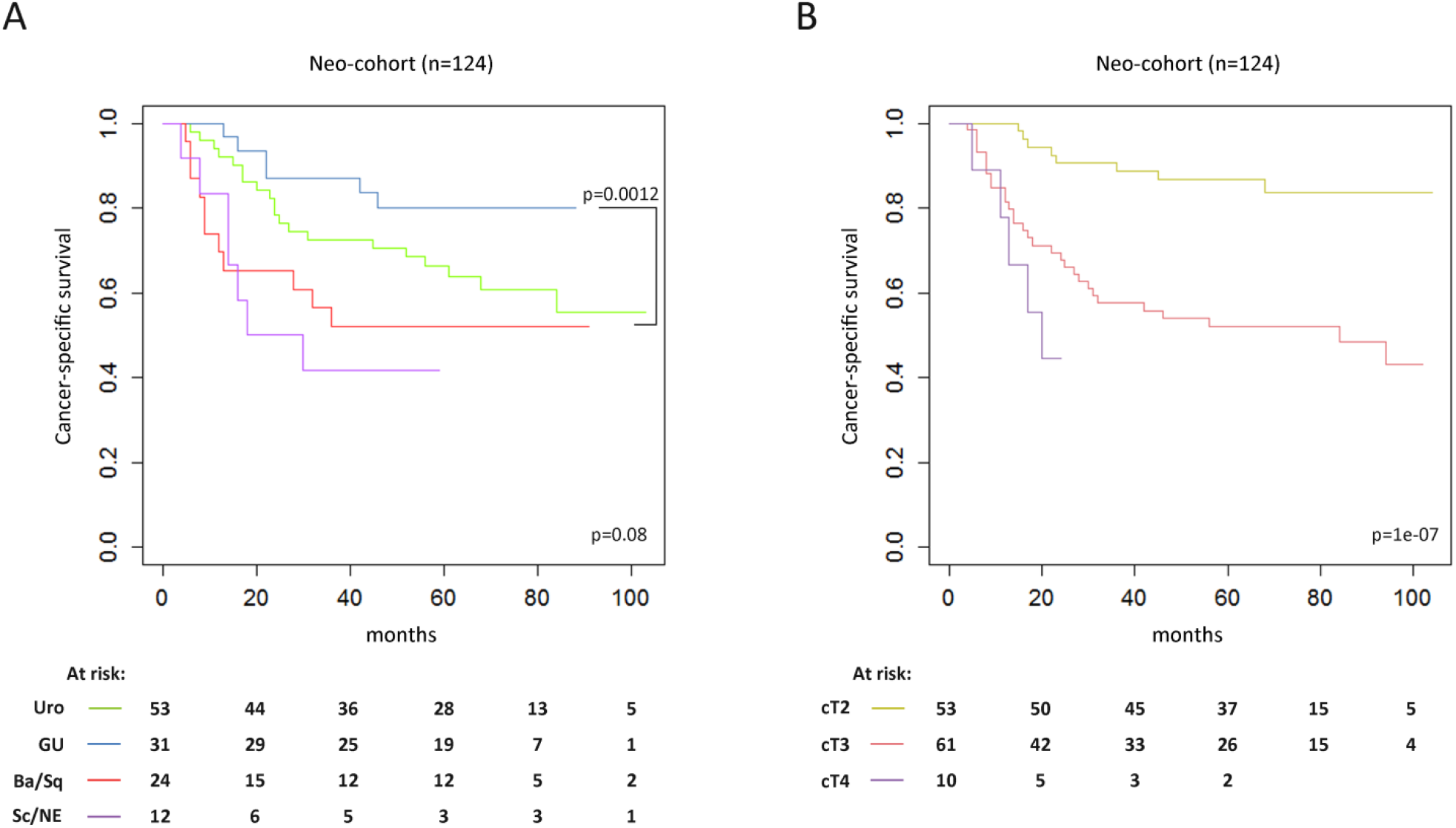
Molecular subtypes and clinical stage are associated with cancer-specific survival in patients treated with neoadjuvant chemotherapy. Kaplan-Meier curves for cancer-specific survival (CSS) stratified by the LundTax subtypes in A), and by clinical stage in B). The Mes-like subtype was excluded due to few patients (n=4). Log-rank p-values are shown including a directed comparison between the GU and Ba/Sq subtypes.

**Table 2.** Multivariable CoxPH analysis of cancer-specific survival including LundTax molecular subtype and clinical stage.

| **Multivariable association to cancer-specific survival (Neo-cohort, n=124)** |  |  |  |
| --- | --- | --- | --- |
| **Variable** | **HR** | **95% CI** | **p-value** |
| LundTax RNA subtype (Ba/Sq reference) |  |  |  |
| UroA | 0.51 | 0.20 - 1.29 | 0.15 |
| UroB | 0.70 | 0.26 - 1.87 | 0.48 |
| UroC | 0.37 | 0.14 - 0.94 | **0.036** |
| GU | 0.29 | 0.11 - 0.79 | **0.015** |
| Mes-like | 0.24 | 0.03 - 1.86 | 0.17 |
| Sc/NE | 1.61 | 0.62 - 4.16 | 0.33 |
| Clinical T-stage (cT3-T4a) (cT2 reference) | 5.76 | 2.63 - 12.59 | **1.16e-05** |

### Validation of pathological response and survival with LundTax IHC subtyping and Consensus classifiers

To validate the observed associations between molecular subtypes and both pathological response and survival in patients treated with neoadjuvant chemotherapy, we also applied the IHC-LundTax subtyping marker panel and the Consensus classification system.^9^ The pCR proportions were similar among subtypes when determined by IHC (GU 42%, Uro 30%, and Ba/Sq 21%) as for the LundTax RNA classifier (GU, 41%; Uro, 32%; Ba/Sq, 21%). Correspondingly, the Lum-Unstable Consensus subtype displayed pCR in 10/19 patients (53%) and the BASQ Consensus subtype in 7/28 patients (25%). Survival analyses in the Neo-cohort stratified by the LundTax IHC classifier using the same multivariable CoxPH regression model as above using CSS, showed similar risk estimates as for the LundTax RNA classifier, with HRs 0.24 (95% CI: 0.09 - 0.60, p=0.003) for GU, 0.36 (95% CI: 0.16 - 0.79, p=0.011) for Uro, and 6.5 (95% CI: 2.86 - 14.77) for clinical stage group cT3-T4a (Supplementary Table 7). Applying the Consensus subtypes in the same model, the HRs were 0.23 (95% CI: 0.07 - 0.80, p=0.02) for the Lum-U Consensus subtype and 0.3 (95% CI: 0.10 - 0.90, p=0.03) for the Stroma-rich Consensus subtype with BASQ as reference (Supplementary Table 8). Thus, GU and the corresponding Consensus subtype Lum-U showed higher proportions of pCR than the Basal/Squamous subtype regardless of classification method used.

### Associations between molecular subtypes, pathological response and survival in external data

To compare the results in the Neo-cohort to external data sets, we used the two largest datasets available; i) the Seiler et al. study^12^ with 190 cases meeting our inclusion criteria (Supplementary Figure 7), and ii) the Taber et al. study^10^ using the 96 cases with RNA-sequencing data (Supplementary Figure 8). While tumors classified as GU in the Seiler cohort had the largest proportion of pCR (47%) among the three major subtypes, it was not significantly different from that of Ba/Sq (44%) (Table 3). When also partial response was considered, the proportions for pCR/pPR response in the Seiler cohort were 56% for Uro, 67% for GU, and 53% for Ba/Sq (GU vs Ba/Sq, p=0.12). However, in the Taber cohort, the proportions of LundTax subtypes with clinical response showed a similar distribution to the proportions with pCR in the current Neo-cohort (UroA, 71%; GU, 67%; Ba/Sq, 40%). Importantly, when all three cohorts were combined, the composite response rates for GU (53%) and UroA (54%) were significantly higher than that of Ba/Sq (38%) (p=0.029 and p=0.031, respectively) (Table 3).

**Table 3.**
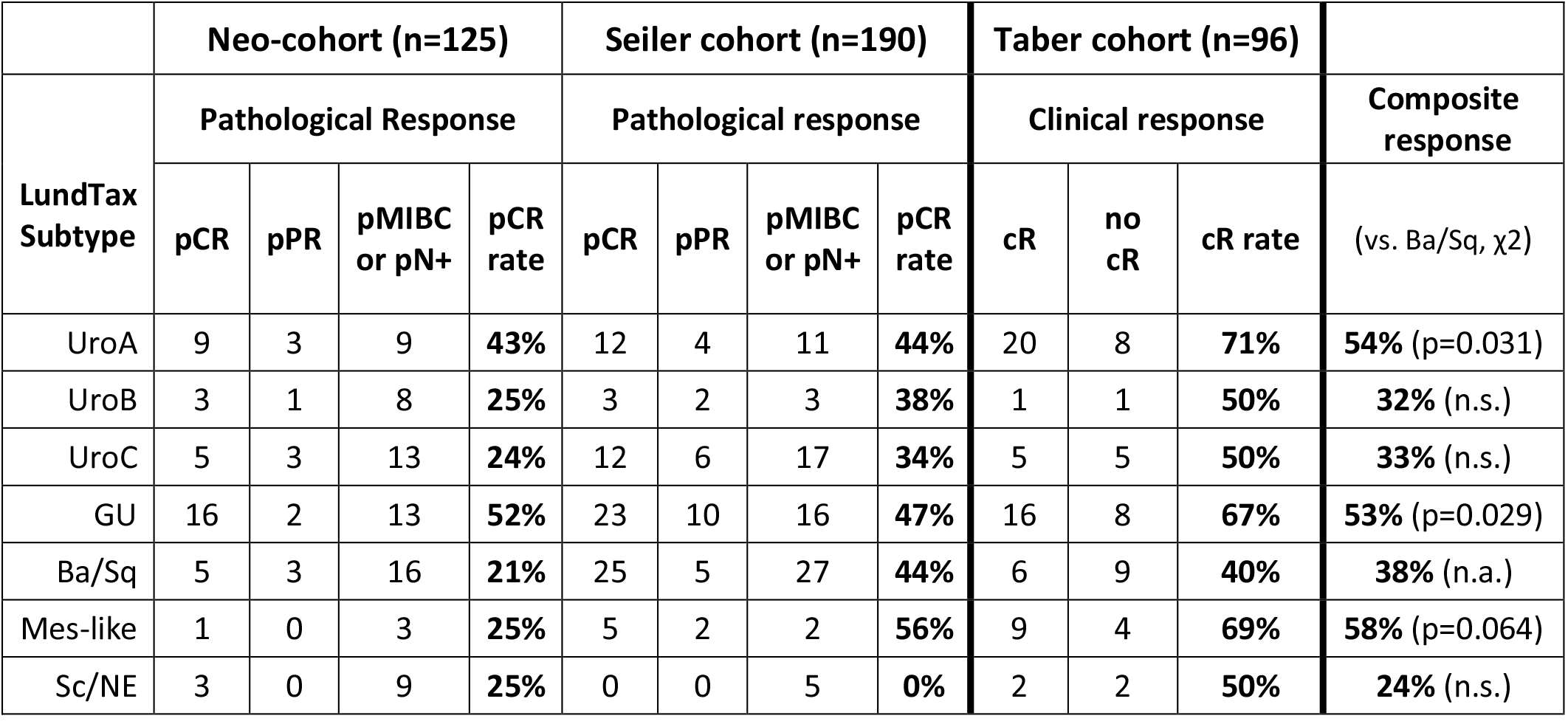
Numbers and proportions of patients with chemotherapy response stratified by LundTax RNA subtype in the current Neo-cohort, in the Seiler and Taber cohorts, and composite proportions across all three cohorts. pCR, pathologic complete response; pPR, pathologic partial response; cR clinical response.

### Transcriptomic analyses of pathological response

To explore if transcriptomic signatures were associated with pCR after preoperative chemotherapy, we organized the tumors by molecular subtype and visualized a number of known coherent signatures that capture important aspects of UC biology in a heatmap (Figure 3A). Biological signatures were associated with LundTax subtypes with the expected expression patterns, but no obvious differential expression was seen in complete responders compared to patients without a complete response. Then, we performed an unsupervised analysis to identify differentially expressed genes (DEGs) between responders and non-responders. The analyses were performed using two methods (SAM and limma), both in the Chemo- and Neo-cohorts, and also separately in the major molecular subsets (Uro, GU, Uro+GU, Ba/Sq). Taken together, few genes were significantly linked to pCR after false discovery rate (FDR) adjustments (Figure 3B). In the Chemo-cohort, three genes (*SPP1, HOXD10*, and *PARP6*) were found to be significantly associated with pCR, all showing increased expression in non-responders. However, it is possible that multiple genes form a biological signature associated with chemotherapy response, without any of them being identified as a significant DEG alone. To test this, we performed gene set enrichment analysis (GSEA) which identified 114 signatures significantly enriched for genes linked to pCR. Since many of these signatures were not originally derived from cancer studies, we grouped them by UC-relevant biological themes and could show that a majority of the signatures (76/114) were involved in proliferation/cell cycle control. Other themes positively linked to response were oxidative phosphorylation, immune/interferon signatures, urothelial differentiation, proteasome components, and MYC-signaling (Figure 3C). The analysis identified fewer signatures linked to non-response, and among them we identified keratinization/EGF signaling and invasion/tumor microenvironment as recurring themes (Figure 3C).

**Figure 3.**
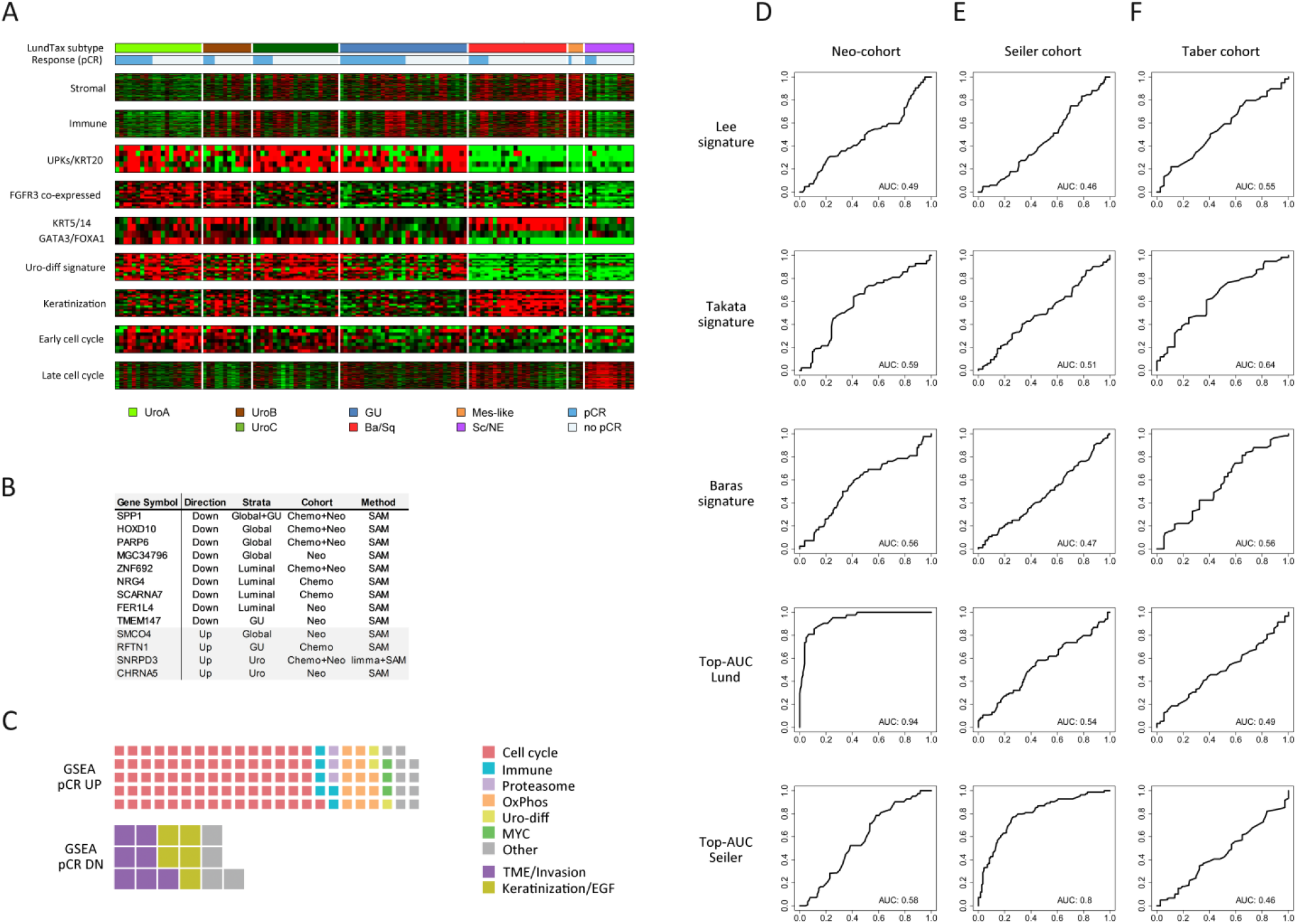
Transcriptomic analyses of pathologic response. Signatures associated with UC biology are shown in A) organized by LundTax subtype and by pathologic response (pCR). The signatures show no apparent global or subtype-specific association with pCR. The heatmap is median-centered on 0 (black) and ranges from -0.9 (green) to +0.9 (red) relative log2 expression. B) Significant differentially expressed genes (DEGs) were identified by SAM/limma in the full Chemo-cohort or Neo-cohort, globally, or stratified into the major LundTax subtypes. *SPP1, HOXD10*, and *PARP6* were the only significant DEGs in the full Chemo-cohort. The wafflechart in C) displays the number of mSigDB signatures annotated into biologically relevant themes that showed positive (GSEA pCR UP) or negative (GSEA pCR DN) enrichment relative to the gene list pre-ranked by the association (limma t statistic) with pathologic response. Panels D)-F) show that previously published and data set specific UC chemotherapy response signatures do not predict response when applied to independent data. D) ROC-curves in the Neo-cohort are shown for the Lee et al., Takata et al., Baras et al., signatures and for simple top-AUC-signatures generated in the Neo-cohort and in the Seiler cohort, respectively. ROC-curves in E) show the same signatures applied to the Seiler cohort. Only the top-AUC-Seiler signatures derived from the same dataset predicts response. Similarly, in F), none of the same signatures applied to the Taber cohort show any predictive capability.

Finally, it is possible that studies with different designs could have identified gene signatures for response that show a signal below what is required for significance in our data. Three published response signatures, Lee,^26^ Takata,^27^ and Baras,^28^ each showed low AUC-values in the Neo-cohort, indicating that none of the signatures were useful for predicting chemotherapy response in this data set (Figure 3D). To further disentangle this lack of predictive capacity, we devised a simple signature (top-AUC-Lund) consisting of the genes in the Neo-cohort that showed top area under the curve (AUC) values. As expected, the mean of this signature performed well in the same data set it was derived from (AUC=0.94), but good performance was also obtained when the analysis was repeated with scrambled (randomized) response labels (AUC=0.88) suggesting that the large AUC was due to overfitting and not a real signal. We then tested the Lee, Takata, and Baras signatures as well as the newly derived top-AUC-Lund signature in the external Seiler cohort. Once again, the three external signatures failed to predict response, as did the top-AUC-Lund signature (AUC=0.54) (Figure 3E). We also derived a similar top AUC-signature from the Seiler cohort and showed that it also performed well in the same data set (AUC=0.80), but not better than with scrambled response labels (AUC=0.90). Finally, when we applied the three external signatures, and the top AUC-signatures derived from the Neo- and Seiler cohorts to the Taber cohort, none of the signatures produced any meaningful chemotherapy response prediction (Figure 3F). Consequently, published signatures for response, as well as our top AUC-Lund signature, are highly cohort dependent and cannot be validated in independent data.

### Osteopontin (SPP1) expression is a subtype dependent response marker

When further investigating the top genes associated with pCR identified above (*HOXD10, SPP1*, and *PARP6*), we found that *HOXD10* and *SPP1* were highly expressed in tumors with Ba/Sq subtype, which may partly explain their link to a poor response. Differential expression among the molecular subtypes could not explain the link between *PARP6* and poor response and this gene may merit further investigating as a potential subtype independent biomarker for chemotherapy response (Figure 4A). On the other hand, *SPP1* coding for osteopontin, a secreted protein widely studied for its role in therapy resistance in cancer, displayed a subtype-dependent effect on response. There was no difference in *SPP1* expression between responders and non-responders within the Uro subtype, but high expression was linked to non-response within each of the other (non-Uro) subtypes (GU, Ba/Sq, Sc/NE, Mes-like) (Figure 4B). We also studied SPP1 expression in the external cohorts used above, and while we found no clear effects in the Seiler cohort (data not shown), we observed similar absence of effect in the Uro subtype in the Taber cohort (Figure 4C). In the Taber cohort the link between high *SPP1* expression and non-response was strongest in the Ba/Sq, Mes-like, and Sc/NE subtypes. To further test *SPP1* expression as a subtype-dependent response marker, we applied an osteopontin antibody to the Chemo-cohort TMAs. The antibody stained tumor cells or stromal cells with a cytoplasmic granular pattern, consistent with its role as a secreted factor (Figure 4D). Additional IHC for alpha-smooth muscle actin and EPCAM were used to facilitate separate evaluation of osteopontin scores in epithelial and stromal compartments. Tumor cell and stromal cell scores correlated only weakly to each other (Pearson r=0.29), but both showed strong positive correlation to the matched relative *SPP1* expression values (Pearson r=0.55 and r=0.46, respectively) (Supplementary Figure 9). We observed that both the tumor and stromal scores were significantly associated with non-response, but only when analyzed within the non-Uro subtypes (Figure 4E). By combining the tumor and stroma scores, the association to response was strengthened but the effect was still limited to the non-Uro subset (Figure 4E). In a multiple logistic regression for pCR adjusting for clinical stage, one unit increase of the combined SPP1 score (ranging from 0-6) was associated with an OR of 0.78 (95% 95% CI: 0.59 - 1.03).

**Figure 4.**
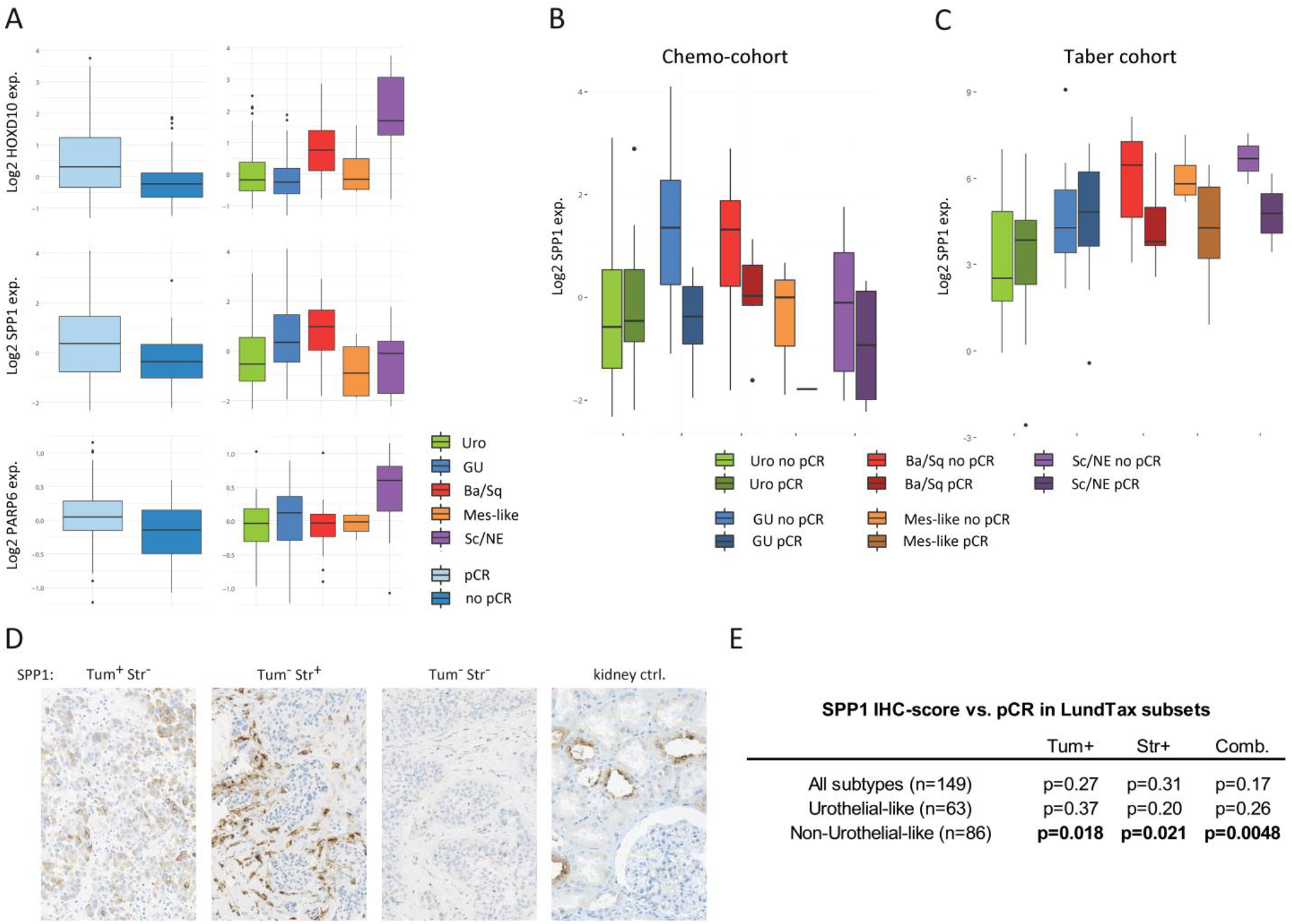
Expression of *SPP1* coding for osteopontin is a biomarker for response only within the non-Uro subtypes. Boxplots in A) show the expression of the three differentially expressed genes (DEGs) stratified by pathologic complete response (pCR) and by LundTax subtypes. *HOXD10* and *SPP1* show increased expression in the poorly responding Ba/Sq subtype. In B), boxplots display the subtype-specific expression of *SPP1* in patients who achieved pCR versus those who did not, within molecular subtypes. Darker colors indicate pathologic complete responders and brighter colors indicate pathologic non-complete responders. Boxplots in C) show the same analysis as in B) i.e. subtype-specific expression of *SPP1* in responders and non-responders in the Taber cohort. Images in D) show 0.24 x 0.36 mm examples of osteopontin (SPP1) IHC positivity in tumor cells and stromal cells and control tissue. The table in E) shows the association between osteopontin (SPP1) IHC-scores and pathologic complete response in the full Chemo-cohort, and in the Uro and non-Uro LundTax subsets separately (Mann Whitney U test).

### Status of UC tumor suppressors has subtype dependent associations to response

UC molecular subtypes are closely linked to inactivating alterations in tumor suppressors p16 (*CDKN2A*), Rb (*RB1*), and p53 (*TP53*). Homozygous deletion of *CDKN2A* or single-copy loss of 9p is frequent in the Uro subtype, can also occur in the Ba/Sq subtype, but is rare in the GU subtype. *TP53* and *RB1* alterations have the opposite pattern, i.e. frequently occurring in GU and Ba/Sq tumors, but rarely in the Uro subtype. We and others have shown that immunostaining for the corresponding proteins can reliably identify these genomic alterations in UC and other tumor types.^22-23^ Thus, the status of these three tumor suppressors was assessed by IHC (Figure 5A). The staining patterns indicative of alteration showed the expected subtype distribution (Supplementary Figure 10). In the Neo-cohort p16 loss by IHC was associated with non-response (p=0.0091), whereas lost or altered Rb or p53 were positively associated with response (p=0.022 and p=0.045, respectively) (Figure 5B). Since the status of these tumor suppressors is closely, but not absolutely, associated with the Uro subtype, we also investigated the response given these alterations and the patient’s molecular subtype. The analyses revealed an association between p16 loss and non-response within Uro (p=0.015), but not within the non-Uro subtypes (p=0.11) (Figure 5C). Conversely, within non-Uro subtypes (GU, Ba/Sq, Sc/NE, Mes-like), nearly all responders (25/26, 96%) had either altered Rb or p53, compared to 35/55 patients (64%) among non-responders (p=0.0018). Altered Rb/p53 was not associated with response in the Uro subtype (p=0.75) (Figure 5D). Finally, we show that while Rb/p53-status did not significantly affect CSS (Figure 5F), patients with Uro subtype and altered p16-status had worse CSS (p=0.02) (Figure 5F). Status of p16 remained significantly associated with CSS in a multivariable CoxPH model including LundTax subtype (Supplementary Table 9). Together with the association to pCR, the survival data indicate that p16 status may be a subtype dependent predictive biomarker for clinical benefit of chemotherapy in patients with the Uro subtype.

**Figure 5.**
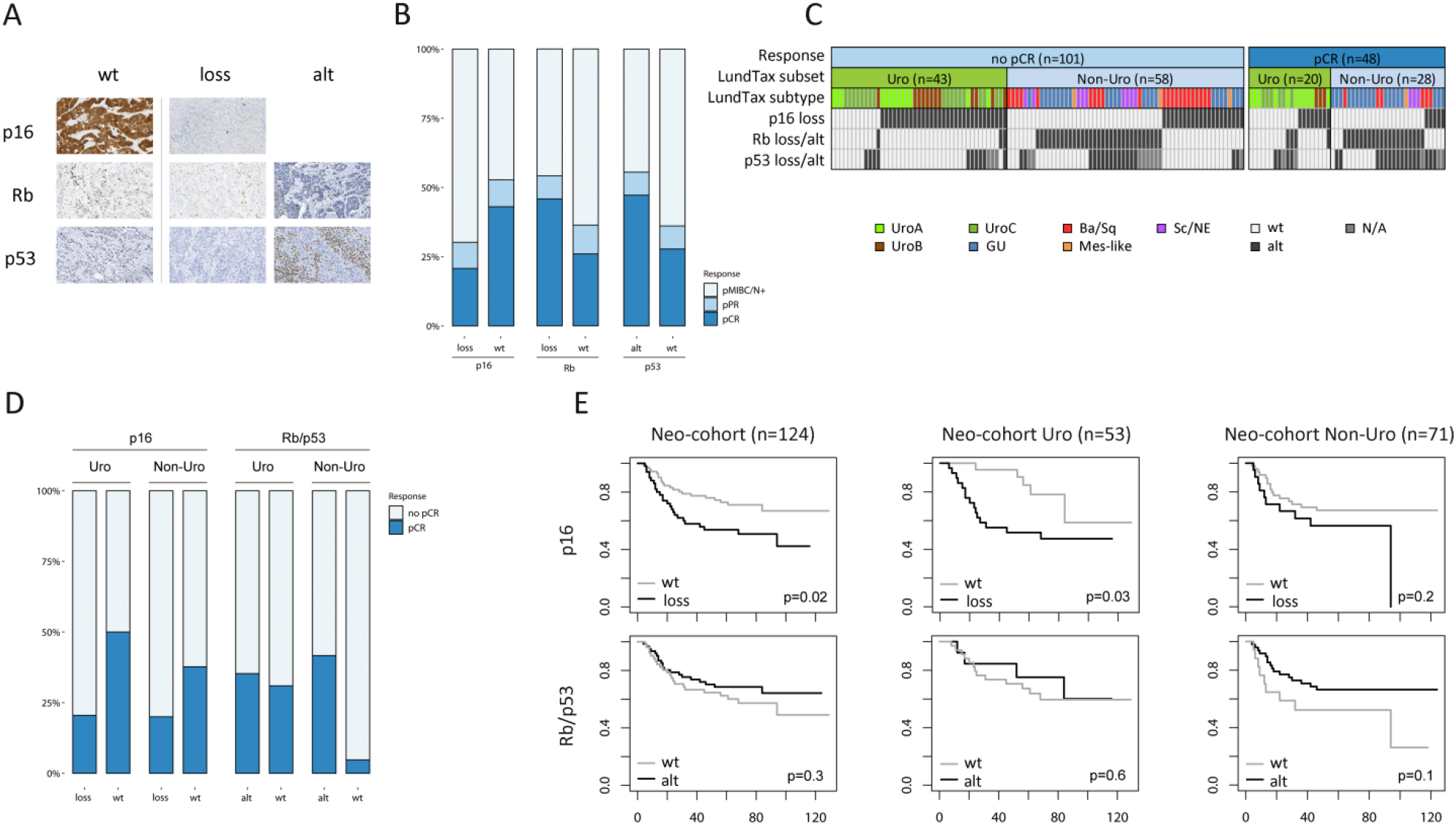
Tumor suppressors p16, Rb, and p53 are associated with chemotherapy response in a subtype-dependent manner. Example images (0.65 x 0.38 mm) are shown in A) of immunostaining in TMA-sections to identify wild-type, loss of, or aberrant expression patterns for p16, Rb, and p53 tumor suppressor proteins. Absent p16 staining indicates *CDKN2A* loss while positive p16 staining indicates wild-type (wt). Absent or perinuclear, Rb staining indicates *RB1* loss or alteration (alt), and absent or overexpressed p53 staining indicates *TP53* loss or alteration. Stacked barplots in B) show the proportions pathologic complete response (pCR) in tumors with altered p16, Rb, and p53 status, respectively. Loss of p16 was associated with poor response, whereas altered Rb or p53 was associated with a better response. The bars and filled boxes in C) show the relationship between pathologic response, molecular subtype and tumor suppressor status. Among patients with a complete response and a non-Uro subtype, all except one had altered status of Rb or p53. In D) the association between p16 loss (left) or altered Rb or p53 status (right) and response is shown separately within the Uro and non-Uro subtypes. Kaplan-Meier plots in E) visualize cancer-specific survival stratified by p16 or Rb/p53 status as above in the full Neo-cohort (n=124) and in the Uro-(n=53) and non-Uro (n=71) subsets.

## Discussion

In this study, we observed improved pathological response and survival outcomes for patients with the GU LundTax molecular subtype independently of tumor stage, following treatment with neoadjuvant chemotherapy and radical cystectomy. Conversely, the Ba/Sq subtype was associated with worse pathological and survival outcomes. The associations to pathological response and survival outcomes were corroborated both by IHC subtyping and by applying the Consensus classification system, by which the LumU subtype (corresponding to GU) also displayed improved pathological response and survival independently of clinical stage compared to the Basal/Squamous subtype. The improved pathological outcome parameters in the neoadjuvant setting was also further validated in an extended cohort comprised of our data set as well as that of Taber et al^10^ and Seiler et al.^12^ In the RC-cohort, treated only with radical cystectomy, the molecular subtypes were not significantly associated to pathological response or survival outcome parameters. Such limited prognostic value for molecular subtypes after radical cystectomy has been reported in previous consecutive and stringently selected cohorts by us^19^ and by other groups,^29^ whereas significant differences in prognosis between molecular subtypes have been reported in cohorts with a wider stage and grade distribution.^8-9, 30^ Based on the findings in the current Neo- and RC-cohorts, we suggest that patients with urothelial cancer of the GU and to some extent Uro subtypes are more responsive to neoadjuvant cisplatin-based chemotherapy than those with Ba/Sq-tumors.

Previous studies using different classification systems to investigate response rates to neoadjuvant chemotherapy across molecular subtypes have not yielded completely consistent results. A meta-analysis based on the Consensus classification system did not reveal any survival differences between subtypes after treatment with neoadjuvant chemotherapy and RC.^9^ However, extrapolated from survival curves it was suggested that the BASQ- and Luminal non-specified (LumNS) subtypes may be more responsive. The present study, applying both LundTax and Consensus classification, found essentially opposite results. Thus the BASQ-subtype consistently displayed both less frequent pCR and worse survival outcomes while the Stroma-rich Consensus subtype showed improved CSS compared to BASQ (HR: 0.3; 95% CI: 0.10 - 0.90). Seiler et al,^12^ applying a different classification system, demonstrated increased overall survival following neoadjuvant chemotherapy in patients with tumors of their basal subtype. Overall survival for this group was increased in comparison with an unmatched patient cohort that did not receive neoadjuvant chemotherapy, but the subtypes were not associated with pathologic response. Notably, most cases that are Ba/Sq according to the Lund taxonomy are termed claudin-low and not basal by their classification.^31^ Thus, the subtype termed claudin-low which had the worst outcome after neoadjuvant chemotherapy and RC in the study by Seiler and colleagues largely corresponds to the LundTax Ba/Sq (Consensus, BASQ) subtype with poor outcome in the current Neo-cohort.

Furthermore, Taber et al,^10^ applying integrated multi-omics analysis in samples from 300 patients receiving either neoadjuvant or first line palliative cisplatin-based chemotherapy, observed worse pathological and radiological outcomes in patients with tumors of the BASQ Consensus subtype, also in line with the findings of the present study. Moreover, the somewhat disparate results in studies investigating neoadjuvant chemotherapy response and molecular subtypes can be attributed to widely varied study designs with different survival endpoints, treatments and selection principles for the cohorts, and possibly also to unknown confounders. For example, recent publications have described how smoking^32^ and secondary MIBC with a previous BCG-treated NMIBC^33^ have worse pathologic response and cancer-specific survival. Such differences are reflected by the varying overall response rates between cohorts: 34% in our population-based study, compared to 42% and 67% in the analyzed subsets of the Seiler et al. and Taber et al. studies, respectively. Beyond cohort differences, the use of different definitions of pathological response (ypT0N0 in the current study, ypTa/pCIS or less in Taber et al,^10^ and ypT<T2N0 in Seiler et al^12^ and ypT<pT1N0 in McConkey et al.^14^) might also contribute. Additionally, the extent of the primary TURB, which is largely unknown in all studies, has impact on the proportion of patients with partial or complete response in the cystectomy specimen, illustrating that survival endpoints are needed in addition to pathologic response to understand differential neoadjuvant chemotherapy response among molecular subtypes. A recent study which classified molecular subtypes by IHC on TMA-cores only, reported improved chemotherapy response and survival in patients with basal/squamous-like tumors.^13^ However, this study included an unknown proportion of carboplatin-based regimens, 17% clinically node-positive patients, and did not report any survival data adjusted for clinical tumor stage. This study used nearly identical markers for the basal/squamous-like subtype as used in the LundTax IHC subtyping. To some extent this suggests that it is not the use of different classification methods but rather actual cohort differences that underlie diverging results between studies.

The need to stratify muscle-invasive UC into subtypes that require different systemic treatment approaches will become ever more important following the introduction of checkpoint inhibitors in the neoadjuvant setting^34^ and new third line regimens for those patients who progress after chemotherapy and checkpoint inhibitors.^35-37^ Based on our results, and the fact that no previous biomarker has reached clinical use, we propose that results in this space may improve with the identification of second generation biomarkers, i.e. those that operate only within a specific context defined by one or several molecular subtypes. The data presented in this study suggest that the bladder cancer relevant^38^ expression of osteopontin (*SPP1*) and well-studied tumor suppressors p16, Rb, and p53 may be such second generation biomarkers that merit further study in this setting. We also note that context-dependency may provide an explanation for the diminishing performance of proposed biomarkers in external validation data sets. In our analyses, three published gene signatures tested in three independent data sets performed only slightly better than chance when used to predict chemotherapy response. The top genes associated with response were different in each data set and tended to only be associated with response in the data set they were identified in. This suggests that identification of global gene signatures for chemotherapy response in bladder cancer is prone to overfitting. It remains to be seen to what extent this situation can be improved by taking molecular subtypes into account in the search for response signatures. It will also be of high relevance to combine gene expression profiling with a more comprehensive genomic analysis in the neoadjuvant setting, including copy-number data, presence of hotspot mutations in *ERCC2*, and other DNA damage response and repair gene alterations which affect cisplatin sensitivity.^10, 16-17, 39^. In a setting of underuse of neoadjuvant chemotherapy^3^ due to fear of overtreatment and toxicity, a possible clinical application of the current findings could be the selection of patients with the GU- and Uro subtypes for such treatment. Conversely, given the likely advent of other neoadjuvant treatment options, it merits further investigation whether patients with the Ba/Sq subtype would be better served by such treatments in place of cisplatin-based chemotherapy.

This study is limited by a small sample size in relation to the number of subtypes, which is reflected in wide confidence intervals for some of the investigated effect measures. In addition, the small number of patients displaying Mes-like and Sc/NE subtypes does not allow for reliable conclusions to be drawn regarding these rare subtypes based on our dataset alone. For clinical implementation of a subtype-diversified use of neoadjuvant chemotherapy to decrease numbers needed to treat, an intention-to-treat approach is also needed, i.e. not excluding patients that only tolerated one chemotherapy course, which is another limitation of the present study. Strengths of the present study include a population-based cohort composition, the employment of both RNA- and IHC-based subtyping as well as the use of two different molecular classification systems and validation in larger combined external datasets.

In conclusion we report that the existing molecular subtypes of MIBC derive different benefit from neoadjuvant chemotherapy as assessed by both pathological response and cancer-specific survival. We suggest that molecular classification and subtype dependent second generation biomarkers should be integrated in future prospective clinical trials with the aim to develop a more precision-based treatment approach for patients with MIBC.

## Data Availability

Gene expression data is available via Gene Expression Omnibus with the accession number GSE169455.

## Acknowledgments

Microarrays were processed at the SCIBLU genomics facility in Lund, Sweden. We thank Per-Ola Bendahl for statistical discussions and Roland Seiler and Peter Black for providing clinical annotations for the Seiler cohort.

## Funding

This work was supported by the Swedish Cancer Society (grant numbers CAN 2017/278 and 2020/0709), Lund Medical Faculty (ALF), The Crafoord Foundation (ref 20130591 and 20140532), Skåne University Hospital Foundation, Torsten Gester Foundation, Gyllenstierna Krapperup’s Foundation (KR2017-0031, KR2018-0050, KR2019-0019), Sten K Johnsson Foundation, Sigurd and Elsa Golje Foundation (LA2017-0222), Magnus Bergvall Foundation, Maud and Birger Gustavsson Foundation, Malmö General Hospital Cancer Foundation, Royal Physiographic Society of Lund, Skåne County Council’s Research and Development Foundation, Gunnar Nilsson Cancer Foundation, Gösta Jönsson Research Foundation, Foundation for Urological Research (Ove and Carin Carlsson bladder cancer donation), and Hillevi Fries Research Fund. The funding sources had no role in the study design, data analyses, interpretation of the results, or writing the manuscript.

## Notes

### Competing Interest Statement

The authors have declared no competing interest.

### Author Declarations

The study was approved by the Research Ethics Board of Lund University (2012/22; 2013/264; 2017/37)

