## Supplementary figures and tables for "Different responses to neoadjuvant chemotherapy in urothelial carcinoma molecular subtypes"

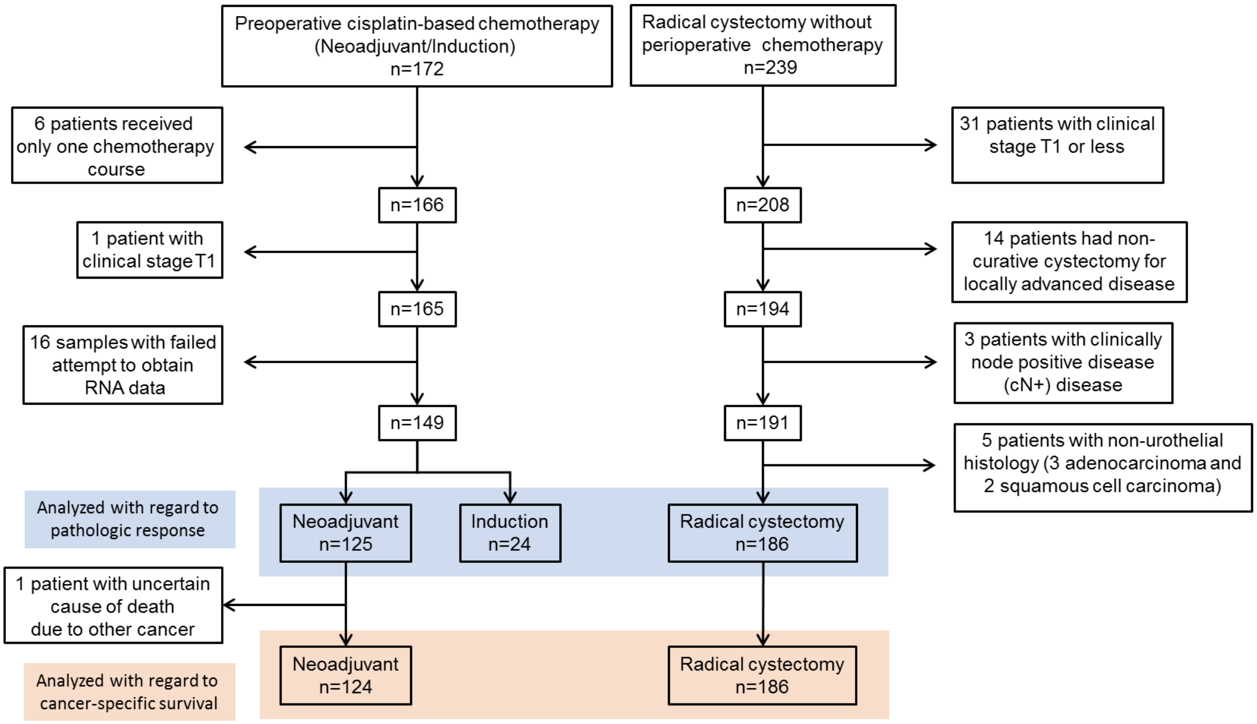


**Supplementary Figure 1**. Flowchart showing the criteria for entry into the Chemo-cohort and in the RC-cohort. From a total of 172 patients treated with cisplatin containing preoperative chemotherapy, 23 were excluded for reasons given in the flowchart. Among 125 patients treated with neoadjuvant therapy, one patient had an uncertain cause of death due to another simultaneously disseminated cancer, leaving 124 patients for survival analyses in this cohort. In the RC-cohort, 239 received radical cystectomy without perioperative chemotherapy. This number was reduced to 186 by excluding patients who received RC for NMIBC, patients operated with non-curative intent, patients with clinically node positive disease, and patients with non-urothelial cancer.

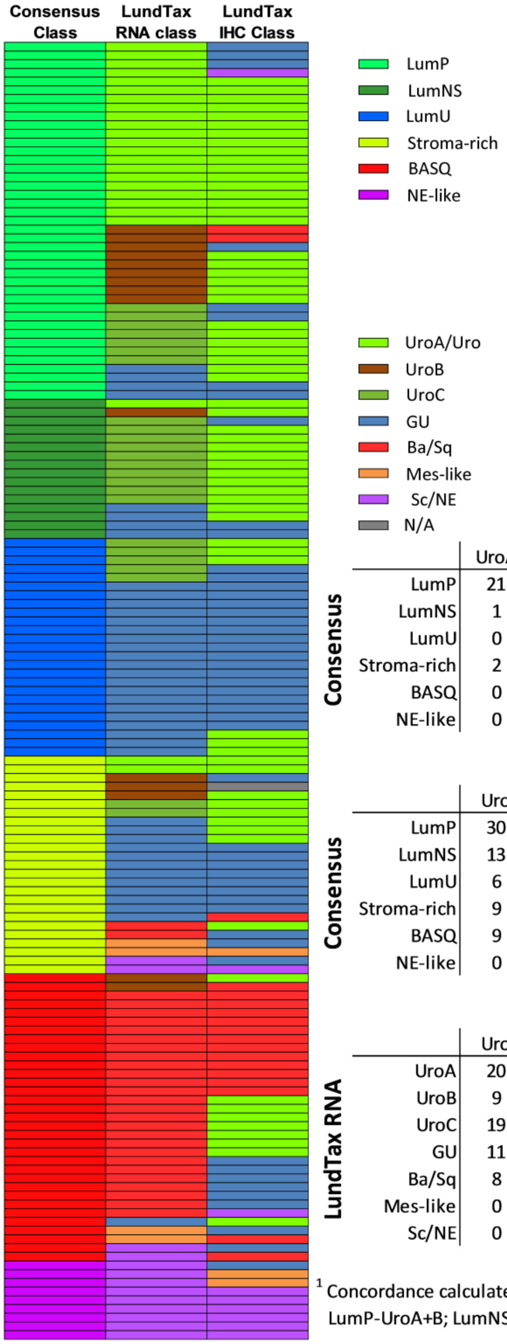


**LundTax RNA**

| Consensus | UroA | UroB | UroC | GU | Ba/Sq | Mes-like | Sc/NE |
|---|---|---|---|---|---|---|---|
| LumP | 21 | 9 | 7 | 4 | 0 | 0 | 0 |
| LumNS | 1 | 1 | 10 | 4 | 0 | 0 | 0 |
| LumU | 0 | 0 | 5 | 20 | 0 | 0 | 0 |
| Stroma-rich | 2 | 3 | 2 | 12 | 2 | 2 | 2 |
| BASQ | 0 | 2 | 0 | 1 | 26 | 2 | 2 |
| NE-like | 0 | 0 | 0 | 0 | 0 | 0 | 9 |

Concordance[1] = 0.77

**LundTax IHC**

| Consensus | Uro | GU | Ba/Sq | Mes-like | Sc/NE | N/A |
|---|---|---|---|---|---|---|
| LumP | 30 | 8 | 2 | 0 | 1 | 0 |
| LumNS | 13 | 3 | 0 | 0 | 0 | 0 |
| LumU | 6 | 19 | 0 | 0 | 0 | 0 |
| Stroma-rich | 9 | 12 | 1 | 1 | 1 | 1 |
| BASQ | 9 | 8 | 15 | 0 | 1 | 0 |
| NE-like | 0 | 1 | 0 | 2 | 6 | 0 |

Concordance[1] = 0.67

**LundTax IHC**

| LundTax RNA | Uro | GU | Ba/Sq | Mes-like | Sc/NE | N/A |
|---|---|---|---|---|---|---|
| UroA | 20 | 3 | 0 | 0 | 1 | 0 |
| UroB | 9 | 2 | 3 | 0 | 0 | 1 |
| UroC | 19 | 5 | 0 | 0 | 0 | 0 |
| GU | 11 | 29 | 1 | 0 | 0 | 0 |
| Ba/Sq | 8 | 7 | 12 | 0 | 1 | 0 |
| Mes-like | 0 | 2 | 1 | 1 | 0 | 0 |
| Sc/NE | 0 | 3 | 1 | 2 | 7 | 0 |

Concordance = 0.66

[1] Concordance calculated by best matching between Consensus and LundTax subtypes as follows: LumP-UroA+B; LumNS-UroC; LumU-GU; Stroma-rich-No matching LundTax subtype; BASQ-Ba/Sq; NE-like-Sc/NE

**Supplementary Figure 2**. Concordance between the Consensus, the LundTax RNA- and the LundTax IHC classifiers.

| Chemo-cohort (n=149) | n | % |
| --- | --- | --- |
| UroA | 24 | 16.1 |
| UroB | 15 | 10.1 |
| UroC | 24 | 16.1 |
| GU | 41 | 27.5 |
| Ba/Sq | 28 | 18.8 |
| Mes-like | 4 | 2.7 |
| Sc/NE | 13 | 8.7 |
| ∑ | 149 | |

| RC-cohort (n=186) | n | % |
| --- | --- | --- |
| UroA | 26 | 14.0 |
| UroB | 16 | 8.6 |
| UroC | 35 | 18.8 |
| GU | 35 | 18.8 |
| Ba/Sq | 50 | 26.9 |
| Mes-like | 13 | 7.0 |
| Sc/NE | 11 | 5.9 |
| ∑ | 186 | |

| Seiler cohort (n=190) | n | % |
| --- | --- | --- |
| UroA | 27 | 14.2 |
| UroB | 8 | 4.2 |
| UroC | 35 | 18.4 |
| GU | 49 | 25.8 |
| Ba/Sq | 57 | 30.0 |
| Mes-like | 9 | 4.7 |
| Sc/NE | 5 | 2.6 |
| ∑ | 190 | |

| Neo-cohort (n=125) | n | % |
| --- | --- | --- |
| UroA | 21 | 16.8 |
| UroB | 12 | 9.6 |
| UroC | 21 | 16.8 |
| GU | 31 | 24.8 |
| Ba/Sq | 24 | 19.2 |
| Mes-like | 4 | 3.2 |
| Sc/NE | 12 | 9.6 |
| ∑ | 125 | |

| Taber cohort (n=96) | n | % |
| --- | --- | --- |
| UroA | 28 | 29.2 |
| UroB | 2 | 2.1 |
| UroC | 10 | 10.4 |
| GU | 24 | 25.0 |
| Ba/Sq | 15 | 15.6 |
| Mes-like | 13 | 13.5 |
| Sc/NE | 4 | 4.2 |
| ∑ | 96 | |

| Ind-cohort (n=24) | n | % |
| --- | --- | --- |
| UroA | 3 | 12.5 |
| UroB | 3 | 12.5 |
| UroC | 3 | 12.5 |
| GU | 10 | 41.7 |
| Ba/Sq | 4 | 16.7 |
| Mes-like | 0 | 0.0 |
| Sc/NE | 1 | 4.2 |
| ∑ | 24 | |

**Supplementary Figure 3**. RNA-based LundTax subtype classification in the Chemo-, Neo-, and Ind-cohorts, as well as the RC-cohort, and the external Seiler and Taber cohorts.

A

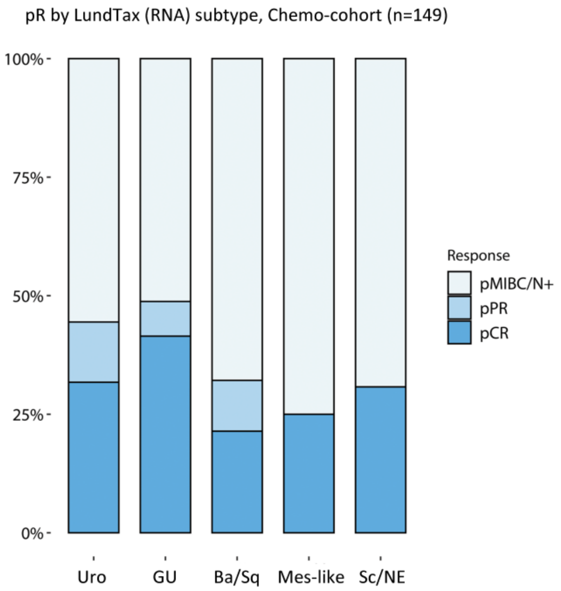


B

pR by LundTax (RNA) classification

| Chemo-cohort (n=149) | pCR | pPR | pMIBC/pN+ | Subtype pCR (%) | Subtype pCR/PR (%) |
|---|---|---|---|---|---|
| Uro (A+B+C) | 20 | 8 | 35 | 32 | 44 |
| UroA | 11 | 3 | 10 | 46 | 58 |
| UroB | 3 | 1 | 11 | 20 | 27 |
| UroC | 6 | 4 | 14 | 25 | 42 |
| GU | 17 | 3 | 21 | 41 | 49 |
| Ba/Sq | 6 | 3 | 19 | 21 | 32 |
| Mes-like | 1 | 0 | 3 | 25 | 25 |
| Sc/NE | 4 | 0 | 9 | 31 | 31 |

| Neo-cohort (n=125) | pCR | pPR | pMIBC/pN+ | Subtype pCR (%) | Subtype pCR/PR (%) |
|---|---|---|---|---|---|
| Uro (A+B+C) | 17 | 7 | 30 | 31 | 44 |
| UroA | 9 | 3 | 9 | 43 | 57 |
| UroB | 3 | 1 | 8 | 25 | 33 |
| UroC | 5 | 3 | 13 | 24 | 38 |
| GU | 16 | 2 | 13 | 52 | 58 |
| Ba/Sq | 5 | 3 | 16 | 21 | 33 |
| Mes-like | 1 | 0 | 3 | 25 | 25 |
| Sc/NE | 3 | 0 | 9 | 25 | 25 |

| Ind-cohort (n=24) | pCR | pPR | pMIBC/pN+ | Subtype pCR (%) | Subtype pCR/PR (%) |
|---|---|---|---|---|---|
| Uro (A+B+C) | 3 | 1 | 5 | 33 | 44 |
| UroA | 2 | 0 | 1 | 67 | 67 |
| UroB | 0 | 0 | 3 | 0 | 0 |
| UroC | 1 | 1 | 1 | 33 | 67 |
| GU | 1 | 1 | 8 | 10 | 20 |
| Ba/Sq | 1 | 0 | 3 | 25 | 25 |
| Mes-like | 0 | 0 | 0 | 0 | 0 |
| Sc/NE | 1 | 0 | 0 | 100 | 100 |

| RC-cohort (n=186) | pCR | pPR | pMIBC/pN+ | Subtype pCR (%) | Subtype pCR/PR (%) |
|---|---|---|---|---|---|
| Uro (A+B+C) | 8 | 18 | 51 | 10 | 34 |
| UroA | 4 | 8 | 14 | 15 | 46 |
| UroB | 1 | 5 | 10 | 6 | 38 |
| UroC | 3 | 5 | 27 | 9 | 23 |
| GU | 4 | 8 | 23 | 11 | 34 |
| Ba/Sq | 2 | 2 | 46 | 4 | 8 |
| Mes-like | 1 | 3 | 9 | 8 | 31 |
| Sc/NE | 0 | 4 | 7 | 0 | 36 |

**Supplementary Figure 4**. A) Pathologic complete response rate stratified by LundTax molecular subtypes in the full chemo cohort (both neoadjuvant and induction treated cases). B) The exact number of cases in each molecular subtype and response category is shown along with calculated percentages of tumors in each subtype that achieved complete response (pCR) or either pCR or partial response (pPR).

A

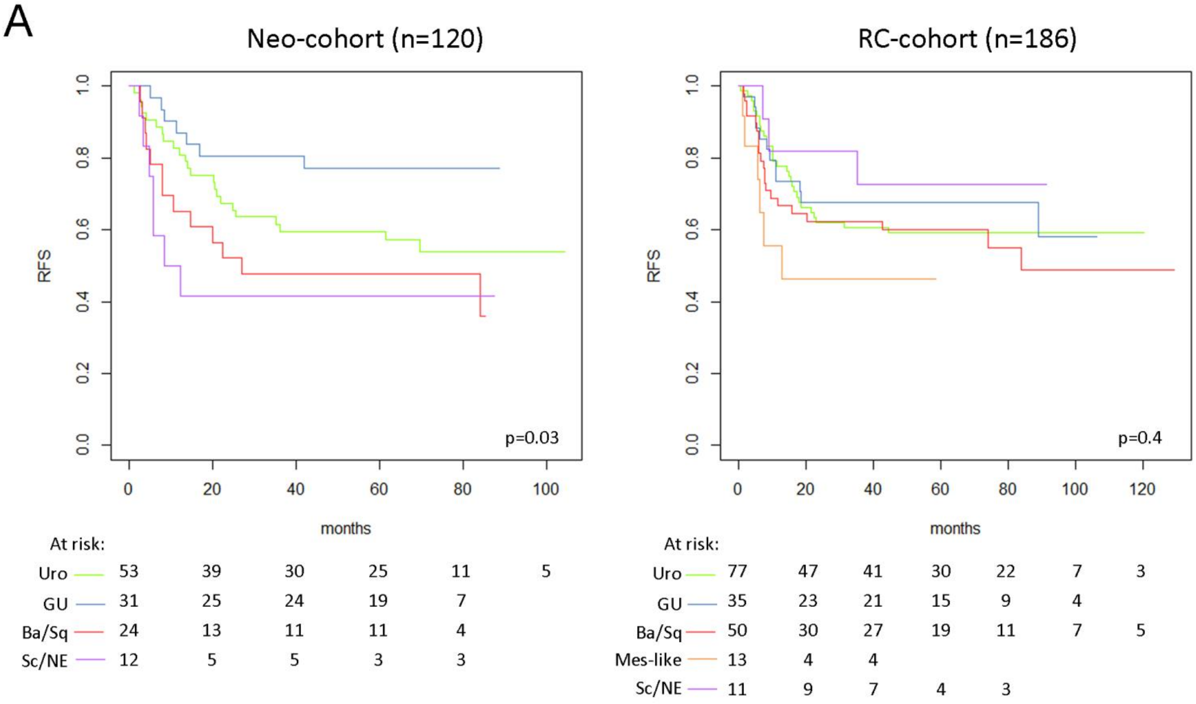


B

**5-year cumulative incidence of recurrence, Neo-cohort**

| Subtype | Recurrence | No recurrence | Censored/followed < 5 years |
|---|---|---|---|
| UroA (n=20) | 8 (53%) | 7 (47%) | 5 |
| UroB (n=12) | 5 (42%) | 7 (58%) | 0 |
| UroC (n=21) | 8 (42%) | 11 (58%) | 2 |
| GU (n=31) | 7 (27%) | 19 (73%) | 5 |
| Ba/Sq (n=24) | 12 (52%) | 11 (48%) | 1 |
| Mes-like (n=4) | 1 (25%) | 3 (75%) | 0 |
| Sc/NE (n=12) | 7 (70%) | 3 (30%) | 2 |

**5-year cumulative incidence of recurrence, RC-cohort**

| Subtype | Recurrence | No recurrence | Censored/followed < 5 years |
|---|---|---|---|
| UroA (n=26) | 12 (52%) | 11 (48%) | 3 |
| UroB (n=16) | 4 (33%) | 8 (67%) | 4 |
| UroC (n=35) | 13 (54%) | 11 (46%) | 11 |
| GU (n=35) | 11 (42%) | 15 (58%) | 9 |
| Ba/Sq (n=50) | 19 (50%) | 19 (50%) | 12 |
| Mes-like (n=13) | 6 (75%) | 2 (25%) | 5 |
| Sc/NE (n=11) | 3 (43%) | 4 (57%) | 4 |

C

**Months to recurrence, Neo-cohort**

| Subtype | 25th %-ile | Median | 75th %-ile |
|---|---|---|---|
| UroA (n=8) | 10.0 | 12.9 | 25.1 |
| UroB (n=7) | 5.6 | 20.3 | 41.3 |
| UroC (n=8) | 6.1 | 14.5 | 20.8 |
| GU (n=7) | 8.1 | 11.5 | 15.4 |
| Ba/Sq (n=13) | 4.3 | 8.1 | 20.0 |
| Mes-like (n=1) | N/A | 7.2 | N/A |
| Sc/NE (n=7) | 4.2 | 5.8 | 7.2 |

**Months to recurrence, RC-cohort**

| Subtype | 25th %-ile | Median | 75th %-ile |
|---|---|---|---|
| UroA (n=12) | 6.5 | 12.7 | 18.1 |
| UroB (n=4) | 6.5 | 9.4 | 13.4 |
| UroC (n=13) | 6.4 | 11.6 | 15.9 |
| GU (n=12) | 5.3 | 9.1 | 12.9 |
| Ba/Sq (n=21) | 5.6 | 7.5 | 11.8 |
| Mes-like (n=6) | 3.0 | 6.2 | 7.4 |
| Sc/NE (n=3) | 8.1 | 9.0 | 22.2 |

D

**Survival after recurrence, Neo-cohort**

| Subtype | Patients alive (Median mo.) | Patients dead (Median mo.) |
|---|---|---|
| UroA (n=8) | - | 8 (3.1) |
| UroB (n=7) | 1 (73.7) | 6 (5.2) |
| UroC (n=8) | 1 (58.5) | 7 (16.8) |
| GU (n=7) | 1 (13.0) | 6 (9.0) |
| Ba/Sq (n=13) | 1 (68.5) | 12 (4.5) |
| Mes-like (n=1) | - | 1 (9.8) |
| Sc/NE (n=7) | - | 7 (8.0) |

**Survival after recurrence, RC-cohort**

| Subtype | Patients alive (Median mo.) | Patients dead (Median mo.) |
|---|---|---|
| UroA (n=21) | 2 (46.7) | 10 (8.2) |
| UroB (n=4) | - | 4 (2.5) |
| UroC (n=13) | 1 (8.6) | 12 (2.4) |
| GU (n=12) | - | 12 (1.9) |
| Ba/Sq (n=21) | 2 (39.9) | 19 (1.6) |
| Mes-like (n=6) | - | 6 (0.5) |
| Sc/NE (n=3) | - | 3 (8.2) |

**Legend on the next page.**

**Supplementary Figure 5**. A) Kaplan-Meier curves for recurrence-free survival (RFS) stratified by LundTax RNA subtype in the Neo- and RC-cohorts. B) Rate of recurrence at 5-years stratified by LundTax RNA subtype in the Neo and RC cohorts. C) Months to recurrence for patients with a recurrence stratified by LundTax RNA subtype in the Neo and RC cohorts. D) Number of patients alive with recurrence at end of follow-up, and median lifetime (months) since recurrence event, and number of patients with a recurrence who died from their disease and the median time from recurrence to death (months) stratified by LundTax RNA subtype in the Neo and RC cohorts.

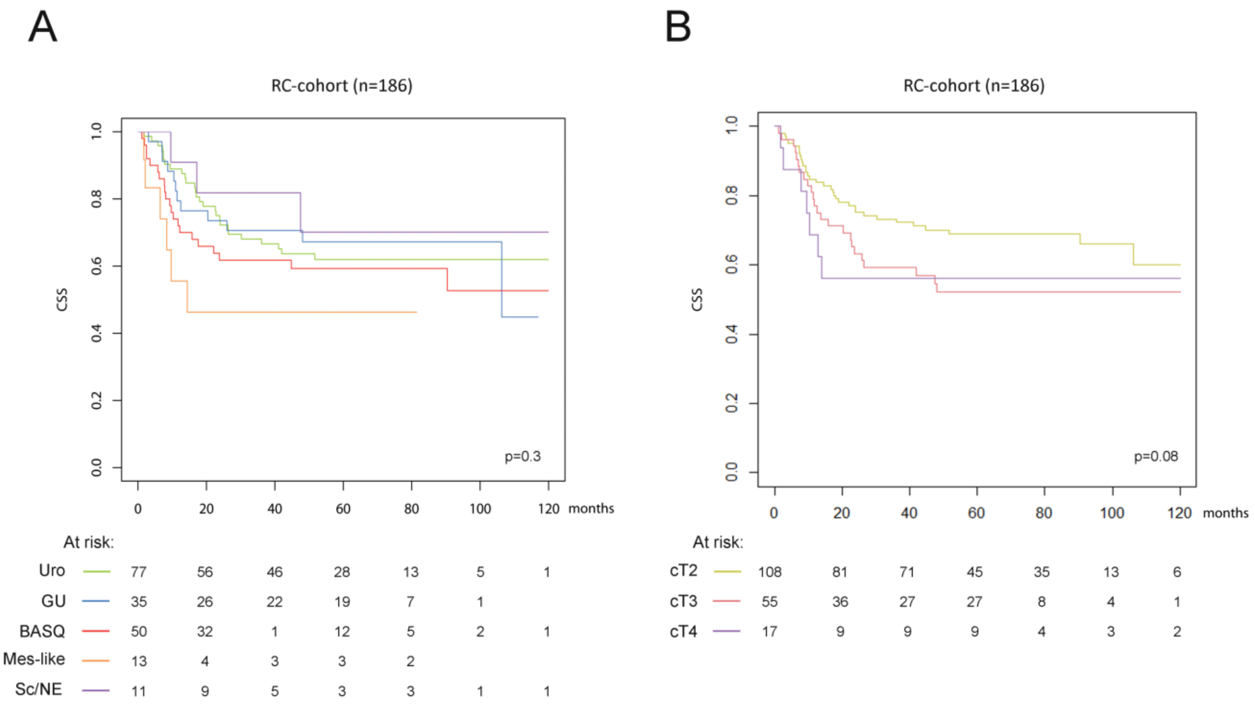


**Supplementary Figure 6**. Kaplan-Meier curves showing the cancer-specific survival (CSS) in the RC-cohort stratified by LundTax molecular subtype (A) or clinical stage (B).

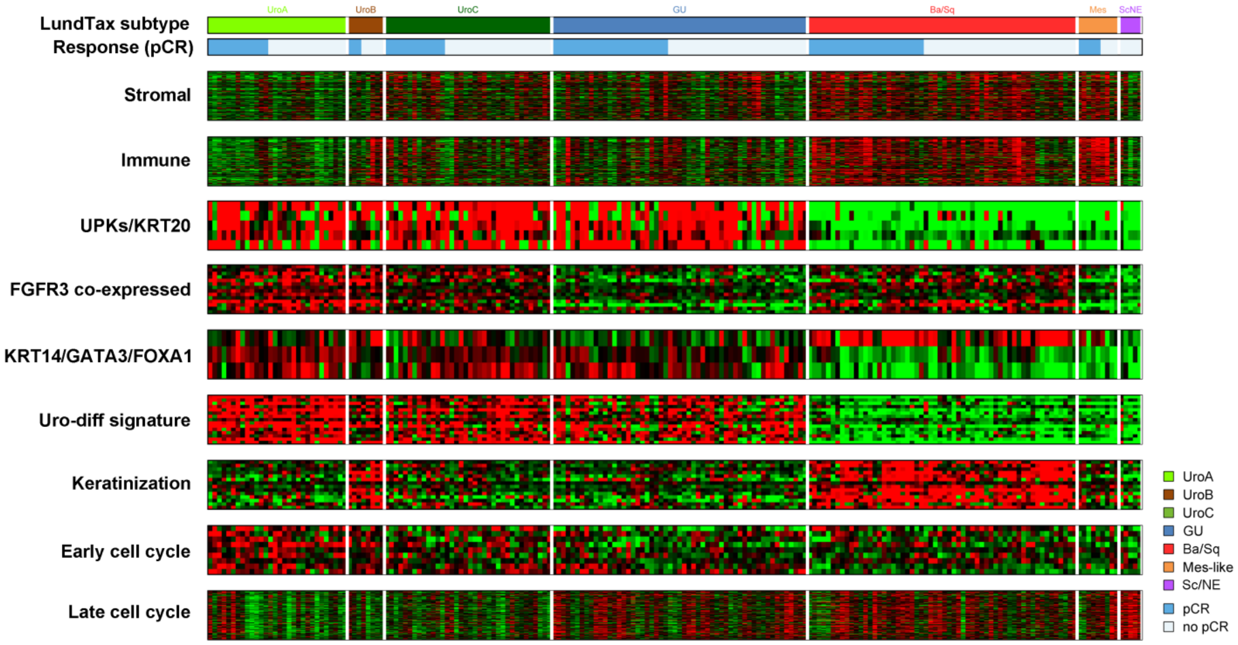


**Supplementary Figure 7**. Distribution of LundTax molecular subtypes, expression of molecular signatures, and pathologic response in the Seiler et al. data set.

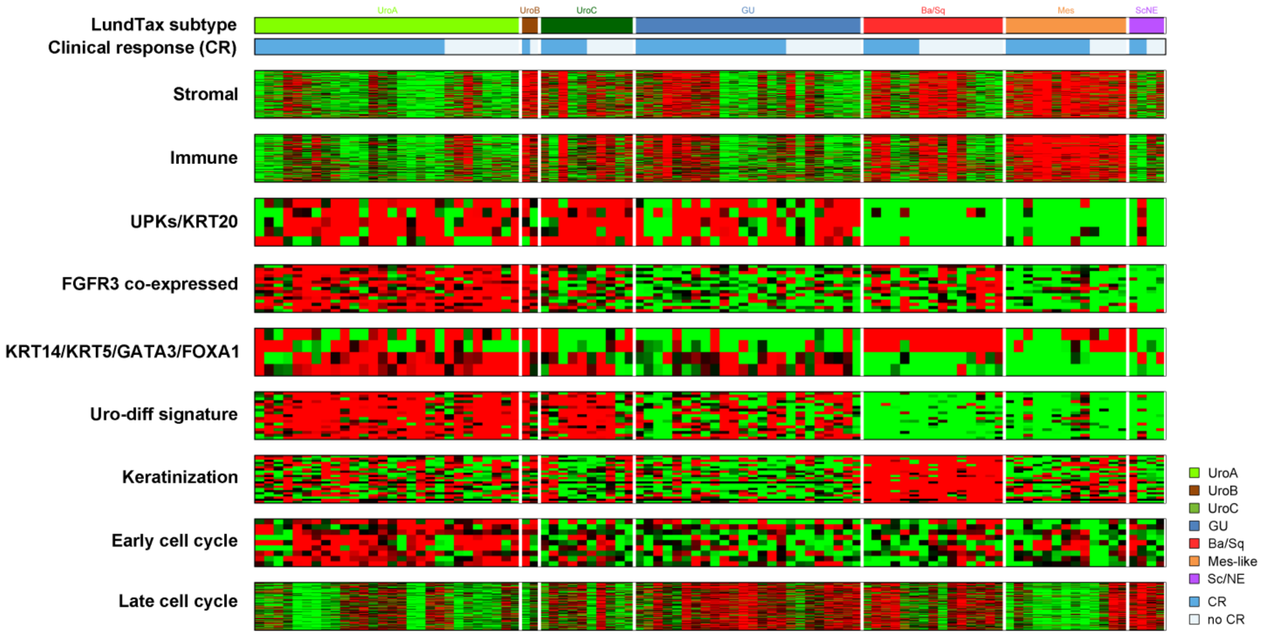


**Supplementary Figure 8**. Distribution of LundTax molecular subtypes, expression of molecular signatures, and clinical response in the Taber et al. data set

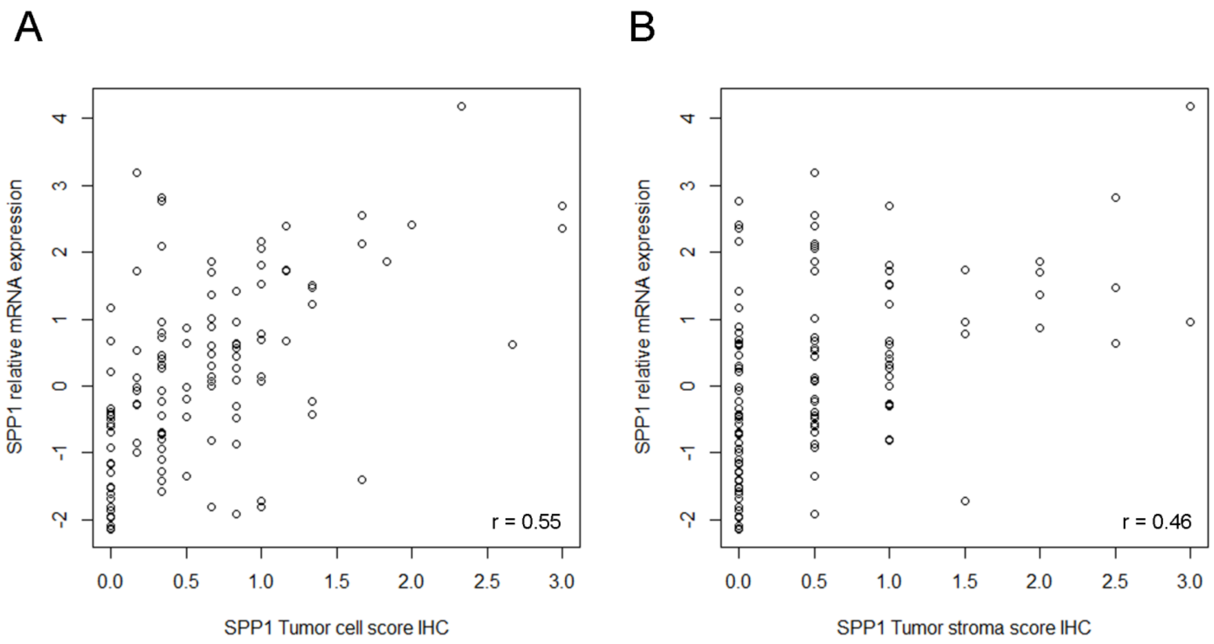


**Supplementary Figure 9**. Scatterplots showing matched data for relative osteopontin (SPP1) mRNA expression versus SPP1 tumor cell IHC-score (A) and stromal cell IHC-score (B).

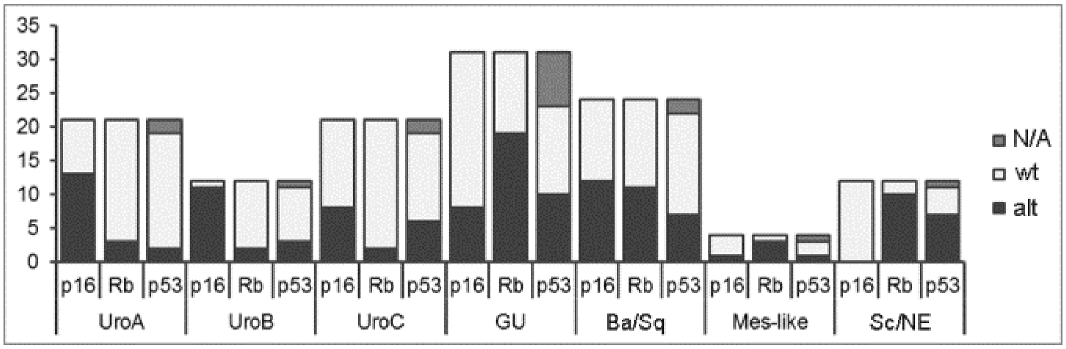


**Supplementary Figure 10**. Barplots displaying the number of tumor suppressor alterations (alt) detected in each LundTax subtype in the Neo-cohort. Uro tumors have high rates of p16 loss, GU tumors have high rates of Rb/p53 alterations, and Ba/Sq tumors have intermediate to high rates of all three tumor suppressors. The Sc/NE subtype has very high rates of Rb and p53 alterations consistent with the established characterization of this tumor type. Note that while both p16 and Rb are markers used to classify UC by IHC, the subtyping used here is LundTax RNA-based classification.

| Markers | Clone | Product number | Vendor | Primary Ab dil. | Evaluation | Comment |
| --- | --- | --- | --- | --- | --- | --- |
| *Basal vs Luminal classification* | | | | | | |
| **KRT5** | EP1601Y | RM-2106 | LabVision | 1:200 | percent, intensity | |
| **KRT14** | LL002 | MS-115 | LabVision | 1:200 | percent, intensity | |
| **FOXA1** | 2F83 | ab40868 | Abcam | 1:200 | intensity | |
| **GATA3** | D13C9 | #5852 | Cell Signaling | 1:800 | percent, intensity | |
| *Genomically Unstable vs Urothelial-like classification* | | | | | | |
| **CCND1** | EP12 | M3635 | Dako (Agilent) | 1:100 | percent, intensity | |
| **FGFR3** | C51F2 | #4574 | Cell Signaling | 1:40 | intensity | |
| **RB1** | 4H1 | #9309 | Cell Signaling | 1:100 | percent | |
| **$p16^{INK4A}$** | | | Ventana | RTU | intensity | |
| *Mesenchymal-like classification* | | | | | | |
| **CDH1** | NCH38 | M3612 | Dako (Agilent) | 1:200 | percent, intensity | |
| **EPCAM** | MOC-31 | M3525 | Dako (Agilent) | 1:40 | percent, intensity | |
| **VIM** | V9 | M0725 | Dako (Agilent) | 1:200 | percent, intensity | |
| **ZEB2** | 6E5 | 61095 | Active motif | 1:500 | intensity | alias SIP1 |
| *Neuroendocrine-like classification* | | | | | | |
| **TUBB2B** | AT5B3 | LS-B4190-50 | LifeSpan | 1:200 | intensity | |
| *Tumor biology markers* | | | | | | |
| **p53** | DO-7 | M7001 | Dako (Agilent) | 1:100 | pattern | |
| **SPP1** | ploycional | HPA027541 | Sigma-Aldrich | 1: 400 | percent (tumor) intensity (stroma) | pos. ctrl. kidney distal tubules |

**Supplementary Table 1**. Antibodies used, staining conditions, and evaluation mode when calculating subtype-scores.

**A**

| Lund-Set signatures | Stromal | Immune | UPKs/KRT20 | FGFR3 co-expressed | KRT5/14/GATA3/FOXA1 | Uro-diff. | Keratinization | Early cell cycle | Late cell cycle |
|---|---|---|---|---|---|---|---|---|---|
| Reference | PMID: 24113773 | PMID: 24113773 | PMID: 29487377 | PMID: 22553347 | PMID: 29487377 | PMID: 26008846 | PMID: 26008846 | PMID: 22553347 | PMID: 22553347 |
| ***Matched genes*** | 97 genes | 115 genes | KRT20 | FGFR3 | KRT5 | BCAS1 | KRT14 | CCND1 | 99 genes |
| | | | UPK1A | TP63 | KRT14 | CYB5A | TGM1 | WEE1 | |
| | | | UPK1B | IRS1 | FOXA1 | CYP4B1 | GSDMC | RBL2 | |
| | | | UPK2 | SEMA4B | GATA3 | CYP4F22 | KRT6A | ID1 | |
| | | | UPK3A | DUOXA1 | | DHRS2 | SFN | ID2 | |
| | | | UPK3B | C16orf74 | | FAM174B | HOXD11 | ID3 | |
| | | | | ZNF385A | | FAM3B | KRT5 | CCNG1 | |
| | | | | SMAD3 | | GGT6 | DSG3 | CCNG2 | |
| | | | | SLC2A9 | | HMGCS2 | KRT6B | | |
| | | | | CLCA4 | | HPGD | HOXD10 | | |
| | | | | SYTL1 | | PPFIBP2 | IL20RB | | |
| | | | | PLCH2 | | PSCA | RHCG | | |
| | | | | SSH3 | | SNCG | AHNAK2 | | |
| | | | | PTPN13 | | TBX3 | SPRR2F | | |
| | | | | DUOX1 | | TRAK1 | FGFBP1 | | |
| | | | | TMPRSS4 | | UPK1A | | | |
| ***Unmatched genes*** | 44 genes | 26 genes | | WNT7B | | VSIG2 | VSNL1 | CDK7 | 47 genes |
| | | | | C3orf54 | | | SERPINB4 | | |
| | | | | CAPNS2 | | | LGALS7 | | |
| | | | | D4S234E | | | SPRR2A | | |
| | | | | | | | BG205162 | | |
| | | | | | | | C12orf54 | | |
| | | | | | | | SPRR2D | | |
| | | | | | | | KRT6C | | |

**B**

| Response signatures | Baras_UP | BARAS_DN | Takata_UP | Takata_DN | Lee_DN |
|---|---|---|---|---|---|
| Reference | PMID: 26230923 | PMID: 26230923 | PMID: 15814643 | PMID: 15814643 | PMID: 17666531 |
| ***Matched genes*** | ZNF486 | HTRA1 | TOP2A | RELA | RHOD |
| | FMO9P | RRAS | MAFB | SLC16A3 | DSP |
| | RHBG | ANKH | DBI | EXT1 | GRB7 |
| | GDPD3 | KLF2 | TCTA | PHKA2 | PPL |
| | SCNN1B | SPRED1 | HMGCS1 | DGKH | SH2D3A |
| | | TFEB | RACGAP1 | | PKP3 |
| | | NRARP | | | HOOK2 |
| | | | | | LLGL2 |
| | | | | | LAD1 |
| | | | | | CDS1 |
| | | | | | CLDN4 |
| | | | | | MYO6 |
| | | | | | MAPK13 |
| ***Unmatched genes*** | ZNF321 | CCPG1 | C14orf142 | | EDG4 |
| | CENTB5 | C9orf125 | RASL11B | | MYO5C |
| | C6orf134 | LOC124220 | PIR51 | | TACSTD1 |
| | LOC400506 | RDHE2 | | | RBM35B |

**Supplementary Table 2**. The Lund set of supervised gene expression signatures (A), and external chemotherapy response signatures (B). Matched genes were used in analyses.

| Multivariable logistic regression analysis of pCR in the Neo-cohort (n=125) | | | | |
|---|---|---|---|---|
| **Variable** | | **odds ratio** | **95% CI OR** | **p-value** |
| LundTax RNA subtype (Ba/Sq ref.) | | | | |
| | GU | 3.46 | 0.95 - 12.59 | 0.059 |
| | Uro | 1.9 | 0.56 - 6.41 | 0.3 |
| | Mes-like | 1.67 | 0.12 - 23.12 | 0.7 |
| | Sc/NE | 1.21 | 0.21 - 6.87 | 0.83 |
| cT (≥T3) | | 0.19 | 0.085 - 0.44 | **0.00011** |

**Supplementary Table 3**. Multivariable logistic regression analysis with pathologic complete response (pCR) as dependent variable. OR, odds ratio; CI, confidence interval; cT, clinical T-stage.

| Variable | | HR | 95% CI | log-rank p | p (Z-dist) |
|---|---|---|---|---|---|
| Sex (male) | | 1.16 | 0.56 - 2.4 | 0.7 | |
| Age at chemostart (year increase) | | 1.012 | 0.97 - 1.05 | 0.6 | |
| Necrosis TURB | | 1.78 | 0.95 - 3.35 | 0.07 | |
| LVI TURB | | 1.16 | 0.62 - 2.15 | 0.6 | |
| Keratinization TURB | | 1.37 | 0.69 - 2.72 | 0.4 | |
| cis TURB | | 1.12 | 0.59 - 2.13 | 0.7 | |
| WHO1999 Grade 3 TURB | | 1.01 | 0.31 - 3.28 | 1 | |
| Histologic variant TURB | | 0.97 | 0.45 - 2.09 | 0.9 | |
| LundTax RNA subtype (Ba/Sq ref.) | | | | 0.08 | |
| | UroA | 0.61 | 0.24 - 1.54 | | 0.29 |
| | UroB | 0.86 | 0.32 - 2.30 | | 0.77 |
| | UroC | 0.49 | 0.19 - 1.25 | | 0.13 |
| | GU | 0.29 | 0.11 - 0.76 | | **0.012** |
| | Mes-like | 0.36 | 0.05 - 2.80 | | 0.33 |
| | Sc/NE | 1.24 | 0.49 - 3.17 | | 0.65 |
| LundTax IHC subtype (Ba/Sq ref.) | | | | **0.002** | |
| | Uro | 0.42 | 0.19 - 0.91 | | **0.03** |
| | GU | 0.21 | 0.09 - 0.55 | | **0.0012** |
| | Mes-like | 0.55 | 0.07 - 4.38 | | 0.57 |
| | Sc/NE | 1.08 | 0.38 - 3.06 | | 0.88 |
| Consensus classification (BASQ ref.) | | | | 0.08 | |
| | LumP | 0.5 | 0.23 - 1.06 | | 0.07 |
| | LumNS | 0.6 | 0.23 - 1.55 | | 0.29 |
| | LumU | 0.22 | 0.06 - 0.76 | | 0.016 |
| | Stroma-rich | 0.31 | 0.10 - 0.94 | | **0.039** |
| | NE-like | 1.99 | 0.78 - 5.17 | | 0.16 |
| cT (≥T3) | | 4.91 | 2.29 - 10.53 | **6E-06** | |
| Regimen type (M-VAC) | | 0.77 | 0.41 - 1.45 | 0.4 | |
| Number of courses | | 0.96 | 0.58 - 1.59 | 0.9 | |

**Supplementary Table 4**. Univariable Cox-proportional hazards (CoxPH) regression for cancer-specific survival (CSS) in the Neo-cohort. HR, Hazard ratio; CI, confidence interval; LVI, lymphovascular invasion; cis, carcinoma in situ; cT, clinical T-stage.

| Multivariable CoxPH analysis of OS in the Neo-cohort (n=124) | | | | |
|---|---|---|---|---|
| **Variable** | **HR** | **95% CI** | **log-rank p** | **p (Z-dist)** |
| | | | **2E-05** | |
| LundTax RNA subtype (Ba/Sq ref.) | | | | |
| UroA | 0.6 | 0.25 - 1.41 | | 0.24 |
| UroB | 0.77 | 0.30 - 1.93 | | 0.57 |
| UroC | 0.44 | 0.19 - 1.04 | | 0.061 |
| GU | 0.41 | 0.17 - 0.96 | | **0.04** |
| Mes-like | 0.23 | 0.03 - 1.77 | | 0.16 |
| Sc/NE | 1.51 | 0.59 - 3.84 | | 0.39 |
| cT (≥T3) | 5.39 | 2.66 - 10.9 | | **2.79E-06** |

**Supplementary Table 5**. Multivariable Cox-proportional hazards (CoxPH) regression for overall survival (OS) in the Neo-cohort. HR, Hazard ratio; CI, confidence interval; cT, clinical T-stage.

| Univariable CoxPH analysis of CSS in the RC-cohort (n=186) | | | | |
|---|---|---|---|---|
| **Variable** | HR | 95% CI | log-rank p | p (Z-dist) |
| Sex (male) | 0.86 | 0.51 - 1.46 | 0.6 | |
| Age at chemostart (year increase) | 1.05 | 1.02 - 1.09 | **0.006** | |
| Necrosis TURB | 1.47 | 0.88 - 2.46 | 0.1 | |
| LVI TURB | 2.56 | 1.58 - 4.15 | **7E-05** | |
| Keratinization TURB | 0.9 | 0.54 - 1.52 | 0.7 | |
| cis TURB | 0.38 | 0.18 - 0.80 | **0.008** | |
| WHO1999 Grade 3 TURB | 1.25 | 0.50 - 3.10 | 0.6 | |
| Histologic variant TURB | 0.6 | 0.22 - 1.65 | 0.3 | |
| LundTax RNA subtype (Ba/Sq ref.) | | | 0.3 | |
| UroA | 0.79 | 0.37 - 1.69 | | 0.55 |
| UroB | 0.55 | 0.19 - 1.60 | | 0.27 |
| UroC | 0.88 | 0.44 - 1.75 | | 0.71 |
| GU | 0.75 | 0.37 - 1.52 | | 0.43 |
| Mes-like | 1.74 | 0.70 - 4.33 | | 0.23 |
| Sc/NE | 0.53 | 0.16 - 1.78 | | 0.3 |
| LundTax IHC subtype (Ba/Sq ref.) | | | 0.2 | |
| Uro | 1.51 | 0.75 - 3.05 | | 0.24 |
| GU | 0.9 | 0.39 - 2.15 | | 0.83 |
| Mes-like | 2.71 | 0.92 - 7.97 | | 0.069 |
| Sc/NE | 0.78 | 0.17 - 3.55 | | 0.75 |
| Consensus classification (BASQ ref.) | | | 0.08 | |
| LumP | 0.81 | 0.41 - 1.58 | | 0.54 |
| LumNS | 0.82 | 0.37 - 1.81 | | 0.62 |
| LumU | 0.66 | 0.30 - 1.47 | | 0.31 |
| Stroma-rich | 1.14 | 0.59 - 2.19 | | **0.7** |
| NE-like | 0.36 | 0.05 - 2.65 | | 0.31 |
| cT (≥T3) | 1.59 | 0.98 - 2.59 | 0.06 | |

**Supplementary Table 6**. Univariable Cox-proportional hazards (CoxPH) regression for cancer-specific survival (CSS) in the RC-cohort. HR, Hazard ratio; CI, confidence interval; LVI, lymphovascular invasion; cis, carcinoma in situ; cT, clinical T-stage.

| Multivariable CoxPH analysis of CSS in the Neo-cohort (n=124) | | | | |
|---|---|---|---|---|
| **Variable** | **HR** | **95% CI** | **log-rank p** | **p (Z-dist)** |
| | | | **2E-07** | |
| LundTax IHC subtype (Ba/Sq ref.) | | | | |
| Uro | 0.36 | 0.16 - 0.79 | | **0.011** |
| GU | 0.24 | 0.09 - 0.60 | | **0.0025** |
| Mes-like | 0.62 | 0.08 - 4.92 | | 0.65 |
| Sc/NE | 2.27 | 0.76 - 6.78 | | 0.14 |
| cT (≥T3) | 6.5 | 2.86 - 14.77 | | **7.77E-06** |

**Supplementary Table 7.** Multivariable Cox-proportional hazards (CoxPH) regression for cancer-specific survival (CSS) in the Neo- cohort using IHC-based LundTax classification. HR, Hazard ratio; CI, confidence interval; cT, clinical T-stage.

| Multivariable CoxPH analysis of CSS in the Neo-cohort (n=124) | | | | |
|---|---|---|---|---|
| **Variable** | **HR** | **95% CI** | **log-rank p** | **p (Z-dist)** |
| | | | **5E-07** | |
| Consensus classification (BASQ ref.) | | | | |
| LumP | 0.47 | 0.22 - 1.01 | | 0.053 |
| LumNS | 0.44 | 0.17 - 1.13 | | 0.089 |
| LumU | 0.23 | 0.07 - 0.80 | | **0.021** |
| Stroma-rich | 0.3 | 0.10 - 0.90 | | **0.032** |
| NE-like | 2.07 | 0.79 - 5.42 | | 0.14 |
| cT (≥T3) | 5.1 | 2.35 - 11.04 | | **3.60E-05** |

**Supplementary Table 8**. Multivariable Cox-proportional hazards (CoxPH) regression for cancer-specific survival (CSS) in the Neo-cohort using Consensus classification. HR, Hazard ratio; CI, confidence interval; cT, clinical T-stage.

| Multivariate CoxPH analysis of CSS in the Neo-cohort (n=124) | | | | | |
|---|---|---|---|---|---|
| **Variable** | | **HR** | **95% CI** | **log-rank p** | **p (Z-dist)** |
| | | | | **0.02** | |
| LundTax RNA subtype (Ba/Sq ref.) | | | | | |
| | UroA | 0.58 | 0.23 - 1.47 | | 0.25 |
| | UroB | 0.62 | 0.23 - 1.70 | | 0.36 |
| | UroC | 0.52 | 0.21 - 1.33 | | 0.17 |
| | GU | 0.34 | 0.13 - 0.91 | | **0.032** |
| | Mes-like | 0.42 | 0.055 - 3.25 | | 0.41 |
| | Sc/NE | 1.94 | 0.69 - 5.46 | | 0.21 |
| p16 loss (IHC) | | 2.31 | 1.16 - 4.63 | | **0.017** |

**Supplementary Table 9**. Multivariable Cox-proportional hazards (CoxPH) regression for cancer-specific survival (CSS) in the Neo-cohort using LundTax RNA classification. HR, Hazard ratio; CI, confidence interval; IHC, immunohistochemistry.
